# The clinical and cost-effectiveness of interventions for preventing continence issues resulting from birth trauma: a rapid review

**DOI:** 10.1101/2024.09.09.24313310

**Authors:** Bethany Fern Anthony, Jacob Davies, Kalpa Pisavadia, Sofie Roberts, Llinos Haf Spencer, Elizabeth Gillen, Juliet Hounsome, Jayne Noyes, Dyfrig Hughes, Deborah Fitzsimmons, Rhiannon Tudor Edwards, Adrian Edwards, Alison Cooper, Ruth Lewis

**Affiliations:** Centre for Health Economics and Medicines Evaluation, Bangor University, United Kingdom; Bangor Institute for Medical and Health Research, Bangor University, United Kingdom; Wales Centre for Evidence Based Care, Cardiff University, United Kingdom; Specialist Unit for Review Evidence, Cardiff University, United Kingdom; Swansea Centre for Health Economics, Bangor University, United Kingdom; Health and Care Research Wales Evidence Centre, Cardiff University, United Kingdom; Health and Care Research Wales Evidence Centre, Bangor University, United Kingdom

## Abstract

Urinary and faecal incontinence, which are often linked to the stresses and strains of childbirth, particularly perineal trauma, are debilitating conditions that can significantly impact womens quality of life. Approximately 85% of vaginal births in the United Kingdom (UK) are affected by childbirth related perineal trauma, either spontaneously or due to episiotomy.

Incontinence also places a significant financial burden on the healthcare system. Previous estimates have shown that stress urinary incontinence alone costs the National Health Service (NHS) 177 million UK pounds per year.

The aim of this rapid review was to identify evidence on the clinical effectiveness and cost-effectiveness of interventions for preventing continence issues resulting from birth trauma. Twenty-three studies, published between 2023 and 2024, were included in this review: 20 systematic reviews of clinical effectiveness and three economic evaluations. A number of key findings, research implications and evidence gaps were identified.

The findings support the use of exercise-based interventions including pelvic floor muscle training for prenatal and postnatal women to prevent urinary incontinence. However, there is limited evidence supporting their long-term effectiveness. Incontinence is a potential long-term burden as pregnancy and childbirth can weaken the pelvic floor, making women more susceptible to incontinence in later life. Menopause often exacerbates these issues due to hormonal changes and by further weakening the pelvic floor muscles. Non exercise-based interventions, such as prenatal perineal massage and vaginal devices were less represented in the available evidence base, especially for faecal incontinence outcomes. There was a paucity of economic evaluations assessing the cost-effectiveness of interventions for incontinence, however, the substantial economic burden of incontinence on the NHS necessitates investment in clinically effective, preventative options. Our findings present the case for investing in exercise-based interventions. Further research is needed to evaluate the maintenance and long-term effects of exercise-based therapy. More research is also needed that focus on alternative type interventions and the prevention of faecal incontinence. Future reviews need to consider qualitative findings of womens experiences and the acceptability and feasibility of rolling out interventions for the prevention of incontinence.

**Funding statement:** The authors and their Institutions were funded for this work by the Health and Care Research Wales Evidence Centre, itself funded by Health and Care Research Wales on behalf of Welsh Government

**EXECUTIVE SUMMARY:** *What is a Rapid Review?:* Our rapid reviews use a variation of the systematic review approach, abbreviating or omitting some components to generate the evidence to inform stakeholders promptly whilst maintaining attention to bias.

*Who is this Rapid Review for?:* This Rapid Review was conducted as part of the Health and Care Research Wales Evidence Centre Work Programme. The review question was suggested by representatives of the Women’s Health Team of the Welsh Government. The intended audience is Women’s Health and continence service commissioners and policy makers in Wales.

*Background / Aim of Rapid Review:* Urinary and faecal incontinence, which are often linked to the stresses and strains of childbirth, particularly perineal trauma, are debilitating conditions that can significantly impact women’s quality of life. Approximately 85% of vaginal births in the United Kingdom (UK) are affected by childbirth related perineal trauma, either spontaneously or due to episiotomy. Incontinence also places a significant financial burden on the healthcare system. Previous estimates have shown that stress urinary incontinence alone costs the National Health Service (NHS) £177 million per year. The prevention of continence issues following childbirth through evidence-based interventions is essential for the health of women both short-term and later in life. The economic cost of incontinence on both individuals and the healthcare system is substantial and the implementation of effective interventions to prevent incontinence following birth trauma can prevent avoidable and costly care in future. The aim of this rapid review was to identify evidence on the clinical effectiveness and cost-effectiveness of interventions for preventing continence issues resulting from birth trauma.

*Results of the Rapid Review:* The evidence base

- The review included evidence available up until June 2024 (when the searches were conducted). The included studies identified in this rapid review were published between 2003 and 2024. Twenty-three studies were included in this rapid review: 20 systematic reviews of clinical effectiveness and three economic evaluations. Key findings

- Twelve systematic reviews of prenatal and/or postnatal pelvic floor muscle training (PFMT) and mixed exercise modes (with a PFMT element) were identified.
- Of the eleven systematic reviews (five of which included meta-analyses) assessing prenatal PFMT and mixed exercise, eight reported findings to support PFMT and exercise for the prevention of urinary incontinence in the postnatal period (up to 6 months postpartum).
- Evidence from two meta-analyses of longer-term outcomes did not support the effectiveness of prenatal PFMT to prevent urinary incontinence in the late postpartum period (defined as greater than 6-12 months) or after 5 years following childbirth. However, data on longer-term outcomes were combined from a diverse set of studies with varied prescription of the PFMT regimens and the reviews did not explicitly examine the impact of continuing PFMT postnatally.
- Of the two systematic reviews that assessed postnatal PFMT one focused on existing incontinence and found no evidence on prevention, and the other found conflicting evidence on the prevention of urinary incontinence.
- Five systematic reviews (of which, three included meta-analyses) explored the effectiveness of prenatal perineal massage. None of the meta-analyses found any significant differences in incidence of urinary incontinence (evidence from three meta-analyses) or faecal incontinence (evidence from two meta-analyses) following prenatal perineal massage. For the other two systematic reviews, one reported a reduction in faecal and gas incontinence but not urinary incontinence, while the other found no effect on any type of incontinence.
- Two systematic reviews reported on the effectiveness of vaginal devices for existing incontinence but did not report on the prevention of incontinence.
- In a systematic review of pushing technique, results demonstrated a significant difference in urinary incontinence scores from baseline to postpartum in the spontaneous pushing group compared with the directed pushing group.
- A cost-utility analysis (conducted to inform NICE guideline 210) utilising a decision analytic Markov model of supervised prenatal pelvic floor muscle training in a population of pregnant women found the intervention to be cost-effective for preventing urinary incontinence when compared to no intervention. The intervention was likely to be cost-effective for all willingness to pay thresholds over £11,000 per QALY gained.
- A cost-effectiveness analysis found that group-based pelvic floor muscle training was more cost-effective than individually supervised training for the prevention of urinary incontinence, at a cost of $14.53 per case of urinary incontinence prevented or cured if eight women attended a training session.
- A RCT and cost-consequence analysis reported no significant difference in urinary or faecal incontinence between groups of nulliparous women adopting an upright or lying down birthing position; but the intervention was not specifically designed to prevent incontinence.

*Policy and Practice Implications:* This rapid review complements existing NICE guidance on the prevention and non-surgical management of pelvic floor dysfunction (NG210, 2021), and the management of faecal incontinence (CG49, 2007). The NICE 2021 guidance recommends pelvic floor muscle training for prenatal and postnatal women, and our rapid review also identified a large evidence base regarding exercise-based interventions to prevent urinary incontinence. However, the NICE guidance indicates limited evidence supporting the long-term effectiveness of PFMT, which also aligns with the findings of our review. We identified two meta-analyses that failed to demonstrate the effectiveness of PFMT in preventing incontinence in the long term, but the included studies varied in prescription of PFMT regimens and did not examine the impact of continuing PFMT postnatally. Incontinence is a potential long-term burden as pregnancy and childbirth can weaken the pelvic floor, making women more susceptible to incontinence in later life. Menopause often exacerbates these issues due to hormonal changes and by further weakening the pelvic floor muscles. Although our review considered a broader range of interventions than the NICE 2021 guidance, other interventions, such as prenatal perineal massage and vaginal devices were less represented in the available evidence base, especially for faecal incontinence outcomes. Despite a paucity of economic evaluations assessing the cost-effectiveness of interventions for incontinence, the substantial economic burden of incontinence on the NHS necessitates investment in clinically effective, preventative options. Our findings present the case for investing in exercise-based interventions. Future recommendations for policy and practice should also consider qualitative findings of women’s experiences and the acceptability and feasibility of rolling out interventions for the prevention of incontinence.

*Research Implications and Evidence Gaps:* A significant evidence gap exists regarding the cost-effectiveness of interventions aimed at preventing incontinence resulting from birth trauma. Further research is needed for non-exercise interventions and maintenance interventions. Future studies adopting longer time horizons are also needed to assess any potential long-term outcomes such as incidence of incontinence during the menopause. Future evidence reviews need to consider qualitative research of the acceptability and feasibility of interventions to prevent continence issues.

## 1. BACKGROUND AND PURPOSE OF THE REVIEW

### 1.1 Who is this review for?

This Rapid Review was conducted as part of the Health and Care Research Wales Evidence Centre Work Programme. The review question was suggested by representatives from the Women’s Health Department of the Welsh Government. The intended audience is Women’s Health and continence service commissioners and policy makers in Wales. This research will be used to guide policy recommendations regarding provisions for continence care in Wales.

### 1.2 Background and purpose of this review

Often linked to the stresses and strains of childbirth, particularly perineal trauma, incontinence is a debilitating condition that can significantly impact women’s quality of life. Perineal trauma and other injuries to this area can weaken pelvic floor muscles resulting in the development of incontinence following childbirth.

Approximately 450,000 women who give birth vaginally each year in the UK suffer some form of childbirth-related perineal trauma. This represents approximately 80% of all vaginal deliveries, either spontaneously, or as a result of an episiotomy (NHS Digital, 2019). Perineal trauma is defined as any damage to the area between the vagina and the anus during childbirth. The severity of trauma is categorised as follows: first-degree (small tears affecting skin that heal without treatment), second-degree (tears affecting perineal muscle typically requiring stitches) and third and fourth-degree tears (deeper tears, also known as obstetric anal sphincter tears, that need to be operated on) (Royal College of Obstetricians and Gynaecologists, n.d.). Third and fourth-degree tears occur in approximately 3% of all vaginal births and 6% of first-time vaginal births in the UK (Royal College of Obstetricians and Gynaecologists, 2015).

The prevalence of perineal trauma varies by mode of delivery. Up to 85% of women who deliver vaginally sustain a form of perineal trauma (NHS Digital, 2019; Smith et al., 2013; Brandie & MacKenzie, 2009). The prevalence of perineal trauma in first time vaginal births (primipara) is estimated to be higher, affecting 91% of women (Smith et al., 2013). Within vaginal births, delivery with instruments (approximately 60 per 1000 deliveries) results in greater incidence of perineal trauma compared to vaginal delivery without instruments (approximately 29 per 1000) (Orlovic et al., 2017). Maternal age, existing maternal medical conditions and a history of complicated deliveries were identified as primary risk factors for perineal trauma (Orlovic et al., 2017).

Incontinence, defined as the accidental loss or leaking of urine, faeces or gas is one of several commonly reported sequelae in women following childbirth. Incontinence can be a significantly debilitating issue in the first year after giving birth (Grant & Currie, 2020; Sobhgol et al., 2022). Estimates from worldwide literature on the incidence of post-partum incontinence in women who delivered vaginally range from 31% to 47% (Moossdorff- Steinhauser et al., 2021; Moran et al., 2020; Thom & Rortveit, 2010).

Urinary incontinence is a devastating condition that can impact women at any stage of the life-course, but is highly prevalent among middle-aged and postmenopausal women, yet it remains a widely underreported issue as many women suffer in silence due to embarrassment or a lack of awareness of treatment options.

Pregnancy and childbirth can weaken the pelvic floor, making women more susceptible to incontinence in later life (NICE, 2021a). Menopause often exacerbates these issues due to hormonal changes and by further weakening the pelvic floor muscles (Menezes et al., 2010). Obstetric trauma from vaginal delivery resulting in irreversible traumatic lesions to the urinary continence system is also a known link to incontinence in later life (Fritel et al., 2012).

The exact prevalence of urinary incontinence among the whole population of women in the UK is varied depending on the definition and methodology of incontinence reported in sources; however, it is estimated that urinary incontinence affects between 20-50% of women, but the prevalence is greater among women who have given birth and in older women (Hannestad et al., 2000; Hunskaar et al., 2004). More recently, a cross-sectional survey of 1,415 women reported a 40% prevalence of urinary incontinence among women in the UK (Cooper et al., 2015). Incontinence (faecal, urinary and gas) are all conditions that reduce quality of life and incur significant costs to both patients and the healthcare system. Implementing preventative interventions targeting incontinence following childbirth has the potential to reduce associated healthcare and wider societal costs. The economic burden of birth trauma and incontinence is presented in section 1.3 of this report.

The initial research question concerned the economic impact of birth trauma. However, following discussions with the stakeholders, the subsequent review question evolved to focus on the cost-effectiveness of interventions to prevent continence issues after birth trauma (or giving birth). The interventions considered were any intervention aiming to prevent continence issues following childbirth.

Preliminary literature searches identified very few economic evaluations of interventions to prevent continence issues following childbirth. To supplement the lack of economic evidence, the scope of the searches for this rapid review was broadened to include evidence from systematic reviews on the clinical effectiveness of interventions to prevent continence issues following childbirth. A separate targeted search of studies reporting economic burden of illness was also conducted to present the economic argument for investing in interventions that have been found to be clinically effective (section 1.3). To further set this review within the relevant UK policy and practice landscape, a presentation of relevant National Institute for Health and Care Excellence (NICE) guidance is presented in section 1.4.

The aim of this rapid review was to present evidence on the clinical effectiveness and cost-effectiveness of interventions to prevent continence issues resulting from birth trauma. The rapid review question was:

> *What is the clinical effectiveness and cost-effectiveness of interventions for preventing continence issues resulting from birth trauma?*

Results from the rapid review search are presented throughout section 2 and grouped by intervention type.

### 1.3 The economic burden of birth trauma and incontinence

A recent UK Government report highlighted the lack of research on the economic cost of birth trauma (The All-Party Parliamentary Group on Birth Trauma, 2024). Identified evidence from the supplementary searches of economic burden focussed on the more immediate costs of perineal trauma (healthcare costs of treating these conditions immediately following delivery). There was no identified evidence on costs and consequences of incontinence following childbirth. However, issues such as psychological distress and trauma following a difficult birth have been found to have long-lasting impacts and cost society £8.1 billion per annum in the UK (Bauer, Tinelli & Knapp, 2022).

The considerable proportion of women who deliver vaginally each year in the UK suffering some form of perineal trauma (80% of all vaginal deliveries) results in increased healthcare costs as mothers are required to remain in hospital for longer to undergo non-elective short-stay procedures. In England, childbirth-related obstetric trauma caused by vaginal delivery is estimated to add an additional 0.5 bed days to patient length of stay post-delivery, before being discharged (Orlovic et al., 2017). The total cost of these additional bed days attributed to obstetric patient safety events was £14.5 million in 2013/2014. Inflated to June 2024 prices*, this figure is now £19.5 million. The average unit cost of non-elective inpatient short-stay following childbirth (in the NHS) in 2013/14 was calculated to be £1,279. Inflating to June 2024* prices, this rises to £1,714 per patient stay (Orlovic et al., 2017). Unit costs represent all healthcare resources used in providing one average short-term inpatient stay targeting and treating obstetric trauma following childbirth.

The delivery method with the highest associated costs is vaginal delivery without instrument. This delivery method results in fewer perineal trauma events than instrument-assisted delivery, yet the volume of unassisted deliveries drives this increase in costs (Orlovic et al., 2017). This finding is supported by the findings of Okeahialam and colleagues (Okeahialam, Sultan & Thakar, 2022).

Obstetric trauma following childbirth affects a substantial number of mothers each year in the UK, and its treatment places excess pressure on the NHS. In 2013/14, the economic burden of all birth-related obstetrical anal sphincter injuries in the UK ranged between £3.7 million (in assisted vaginal births) and £9.8 million (in spontaneous vaginal births) (Orlovic et al., 2017). Inflated to June 2024 prices*, these figures are £4.9 million and £13.1 million, respectively. From a wider perspective, £3.1 billion was awarded in patient legal damage claims against perineal trauma between 2002 and 2012. It was the fourth leading cause of claims in obstetrics (Steen & Diaz, 2018).

Incontinence, the involuntary loss of urine or faeces can be a significant complication of perineal trauma and childbirth more generally. The economic cost of incontinence on both individuals and the healthcare system is substantial (Javanbakht et al., 2020). Costs arise from various sources, including direct healthcare costs, patient out-of-pocket expenses for incontinence products and wider societal costs such as lost productivity due to absenteeism or reduced work performance (Fultz et al., 2005). A previous study utilising data from three European countries estimated that the total cost of stress urinary incontinence in the UK is approximately £818 million (Papanicolaou et al., 2005). A separate study reported a cost of £118million per year to the NHS in healthcare costs for stress urinary incontinence (Turner et al., 2004).

The prevention of birth trauma and subsequent urinary and faecal incontinence could facilitate immediate cost savings to the healthcare system by reducing time spent in hospital after birth. Interventions to prevent incontinence following childbirth will also yield substantial long-term savings through decreased health and care resource utilisation. Cost-savings from incontinence prevention from a wider societal perspective may be manifested through reduced absenteeism and improved productivity among affected workers.

### 1.4 NICE guidance on perineal care

This section summarises the National Institute for Health and Care Excellence (NICE) 2021 guidance (NG210) on the prevention and non-surgical management of pelvic floor dysfunction (NICE, 2021a) and the NICE 2007 guidance (CG49) on faecal incontinence management in adults (NICE, 2007). The NICE guidance presented in this section will be revisited and considered in relation to the main findings of this rapid review later in the discussion section of this report (Section 3).

The NICE 2021 (NG210) guideline focuses on the prevention and non-surgical management of pelvic floor dysfunction, and cites urinary incontinence, faecal incontinence and pelvic floor prolapse as the three most common and definable symptoms of this condition (NICE, 2021a). The guidance document reports that being over the age of 30 when giving birth, and having previous childbirths are risk factors for pelvic floor dysfunction. Labour-related risk factors include assisted vaginal births (forceps or vacuum), occipito-posterior births (where the baby is lying face-up), an active second stage of labour lasting more than 1 hour, and trauma/injury to the anal sphincter during childbirth (NICE, 2021a). While the issue of incontinence is a significant part of pelvic floor dysfunction, the guideline provides a broader approach to managing these issues. The guidance emphasises prevention by reducing modifiable risk factors which can contribute to incontinence and recommends thorough clinical assessment to determine the type and severity of incontinence (NICE, 2021a).

NICE recommends pelvic floor muscle training (PFMT) for prenatal and postnatal women to prevent symptoms of pelvic floor dysfunction (NICE, 2021a). NICE has recommended PFMT as a first-line treatment for urinary incontinence since 2006 and points to the clinical effectiveness and cost-effectiveness of PFMT as a preventative measure. The 2021 NICE guidance recommends that PFMT should be supervised by a physiotherapist. In the NHS Long Term Plan, physiotherapy was reported as the most cost-effective intervention for preventing and treating mild to moderate incontinence and prolapse (NHS, 2019). The evidence review (NICE, 2012b) supporting the NICE guidance found that PFMT improves several symptoms of pelvic floor dysfunction (pelvic organ prolapse, stress and mixed urinary incontinence, and faecal incontinence with coexisting pelvic organ prolapse).

In France, all postnatal women receive pelvic floor rehabilitation. NICE guidance states that there is not enough strong evidence on the cost-effectiveness of this model for adoption in the UK and makes a recommendation for research on whether universal postnatal PFMT is effective in preventing pelvic floor dysfunction. NICE guidance states that antenatal supervised PFMT is likely to be cost-effective for some pregnant women, in particular women in the groups identified at higher risk of developing pelvic floor dysfunction. The guidance recommends PFMT as an option as it is likely to be cost-effective for some women in these groups. The NICE urinary incontinence quality standard states that pregnant women with stress urinary incontinence or mixed urinary incontinence should be offered a programme of supervised PFMT for at least three months (NICE 2021c). The same recommendation of a three-month supervised programme should also be offered from week 20 of pregnancy to women who have a first-degree relative with pelvic floor dysfunction (NICE 2021a).

Based on the findings of the NICE evidence review, the committee’s discussion of the evidence recommended that all women should be encouraged to do pelvic floor muscle exercises (NICE, 2021b). The 2021 NICE guidance also takes into account recommendations made by the Independent Medicines and Medical Devices Safety Review, which emphasises the life-course model, recommending that pelvic floor education should be encouraged in antenatal classes and in schools, where appropriate (UK Government, 2020). Despite limited evidence on long-term effectiveness, in the committee’s experience continuing with PFMT is key for continued prevention of symptoms, and they agree that low long-term adherence is likely to explain the limited evidence for long-term effectiveness. Recognising these problems with adherence, the committee agree that women should be encouraged to continue PFMT throughout their life. One study that informed the 2021 NICE guidance showed that PFMT significantly reduced the number of post-menopausal women developing urinary incontinence. From this study, NICE recommends further research into whether PFMT is effective in preventing pelvic floor dysfunction for older women (aged 65 and over), and women in the perimenopausal or postmenopausal phases.

The NICE 2021 guidance also highlights that the effectiveness of intravaginal devices when combined with PFMT is still unclear based on current evidence and consequently the committee made a recommendation for research on the effectiveness of vaginal devices (NICE, 2021a).

The NICE 2007 guidance (CG49) emphasises the importance of PFMT as a first-line treatment for obstetric-related faecal incontinence, which is a debilitating condition that may present early after giving birth (NICE, 2007). The guidance highlights that previous birth trauma is a significant cause of faecal incontinence in later life and advocates the value of preventative measures that will benefit both younger and older individuals (NICE, 2007).

NICE advocates that early intervention in the postpartum period may decrease the risk of delayed-onset faecal incontinence in women; however, evidence is lacking on whether prenatal interventions (before obstetric trauma or injury) can provide a protective role in preventing faecal incontinence (NICE, 2007).

## 2. RESULTS

### 2.1 Overview of the Evidence Base

This rapid review aims to complement the NICE 2021 guidance (NG210) on the prevention and non-surgical management of pelvic floor dysfunction, and the NICE 2007 guidance (CG49) on the management of faecal incontinence in adults which was summarised in section 1.4 of this rapid review report. The rapid review findings reported in this section (Section 2) will be considered in relation to the NICE guidance in the discussion section of this report (Section 3). Evidence of clinical effectiveness from systematic reviews studies is presented in Section 2.2, followed by cost-effectiveness evidence from economic evaluations in Section 2.3.

The methods and eligibility criteria used for conducting the review are presented in Section 5. The rapid review search strategy is presented in Appendix 1. After the removal of duplicates, the database searches identified 3,383 references (see Figure 1 in Section 6.1 for the PRISMA diagram). Following title and abstract screening, 56 papers were retrieved for full text screening. Twenty-three studies were included in this rapid review: three economic evaluations (Bick et al., 2017; Brennen et al., 2021; NICE, 2021), and 20 systematic reviews of clinical effectiveness, of which two were reviews of existing reviews (Ryhtä et al., 2023; Sananès et al., 2023). The overlap of primary studies within this rapid review is low at <5%, as shown in the diagram in Appendix 4. However, this assessment of overlap did not include the two reviews of existing reviews (Sananès et al., 2023; Ryhtä et al., 2023). It is important to note that one of the reviews presented in the review of existing Cochrane systematic reviews (Sananès et al., 2023) also included one of the systematic reviews included in this rapid review (Woodley et al., 2020). Of the nine systematic reviews included in the second review of existing reviews (Ryhtä et al., 2023), five are included in this rapid review.

**Figure 1.**
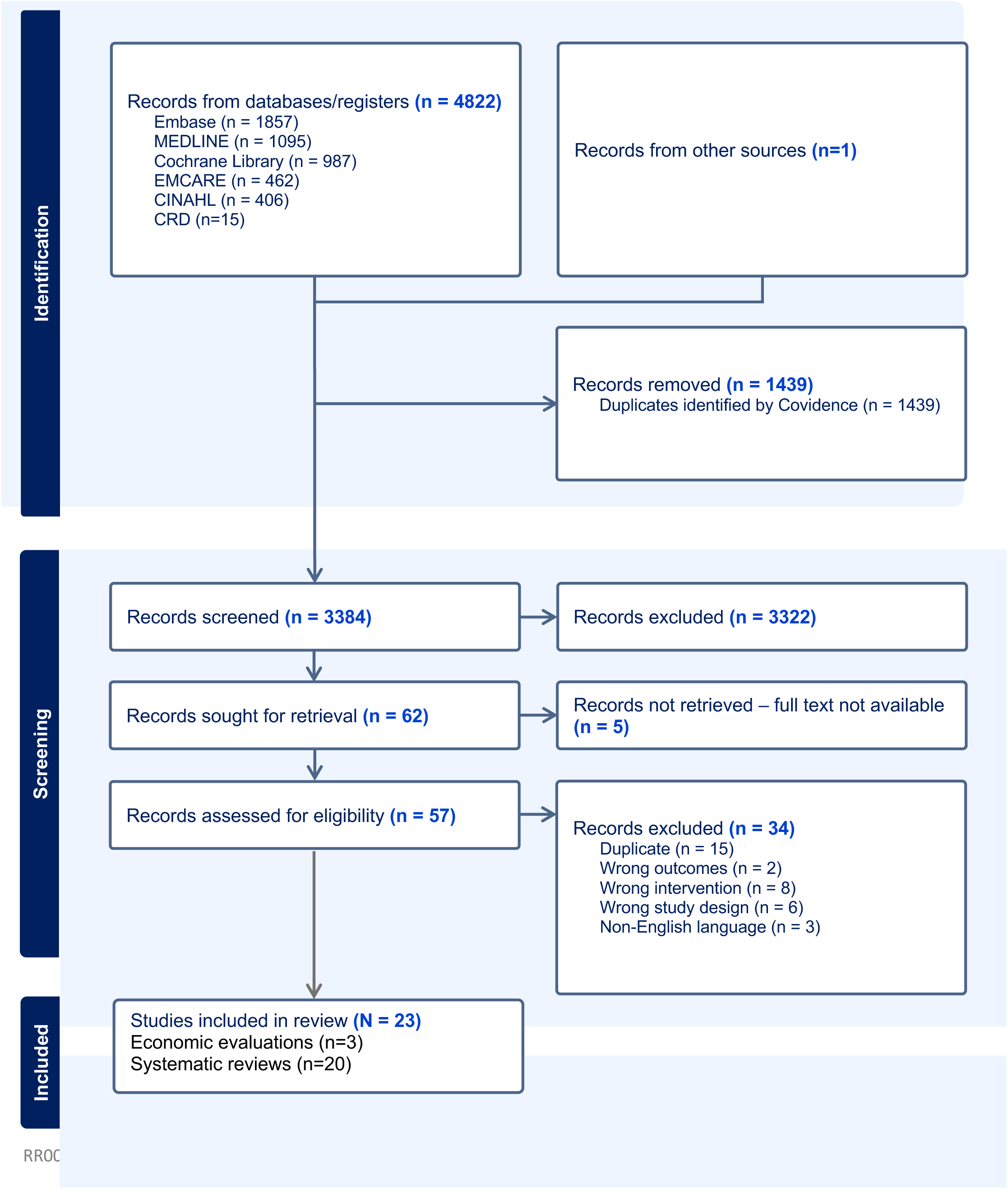
PRISMA 2020 flow diagram of included records (Page et al., 2021)

The 20 systematic reviews included in this rapid review were conducted by authors in the UK (Oblasser et al., 2015; Wagg & Bunn, 2007), Australia (Haddow et al., 2005), Canada (Davenport et al., 2018; Harvey, 2003), New Zealand (Woodley et al., 2020), Belgium (Van Kampen et al., 2015), Switzerland (Sananès et al. 2023), Spain (Perales et al., 2016; Zhang et al., 2023), Finland (Ryhtä et al., 2023), Poland (Milka et al., 2023), Columbia (Mantilla Toloza et al., 2024), China (Chen et al., 2022), Egypt (Abdelhakim et al., 2020), Japan (Shinozaki et al., 2023), Iran (Khorasani et al., 2020), and Brazil (Lemos et al., 2008; Schreiner et al., 2018; Santos et al., 2024). All of the systematic reviews reported on the impact of interventions to reduce pregnancy or birth-related incontinence, with some reviews assessing the effectiveness of multiple interventions. Of the 20 systematic reviews of clinical effectiveness included in this rapid review, eight systematic reviews reported on the effectiveness of PFMT/pelvic floor muscle exercise, four explored mixed exercise interventions, five reviews assessed prenatal perineal massage, one reported on birthing position, and three assessed vaginal devices.

Three economic evaluation studies were included as part of this rapid review. The first study was conducted in England and Wales and included two economic evaluations, a cost-consequence analysis and a cost-effectiveness analysis comparing an upright birthing position with a lying down birthing position (Bick et al., 2017). The second economic evaluation was a cost-effectiveness analysis based on a systematic review of different models of supervised pelvic floor muscle training (PFMT) and was conducted by a team of researchers from Australia (Brennen et al., 2021). The third economic evaluation was a model-based cost-utility analysis of supervised prenatal PFMT utilising existing data from two studies (both reporting on the same supervised PFMT intervention at two separate time points) conducted as part of 2021 NICE guidance (NICE, 2021b).

### 2.2 Effectiveness of interventions to prevent incontinence and birth trauma outcomes

A summary of the findings from the 20 systematic review papers is presented with a specific focus on incontinence outcomes. Intermediate outcomes reported in the systematic reviews that can impact on/or are impacted by incontinence such as birth trauma outcomes, pelvic floor strength, quality of life, and adverse outcomes (e.g., perineal pain, vaginitis) are also presented. The outcomes reported in each systematic review are presented in Table 2 under Section 6.2 of this report. Where possible, information on the type of birthing mode has been presented if information was available in the systematic review papers (Table 2, Section 6.2). A graphical representation of the overlap of primary studies across the systematic reviews is presented in Appendix 4 (Section 8.4). This ‘GROOVE’ assessment of overlap (Bracchiglione et al., 2022) excluded the two reviews of existing reviews as they did not review primary studies. The GROOVE exercise reported a slight overlap (3.5%) across 18 systematic reviews included in this study (Appendix 4, Section 8.4).

The following sub-sections are divided into intervention types reported in the identified SR papers and include the following: pelvic floor muscle training/exercise, mixed exercise programmes, prenatal perineal massage, and birthing position and vaginal devices.

#### 2.2.1 Prenatal and postnatal pelvic floor muscle training

Eight systematic reviews reported on the effectiveness of pelvic floor muscle training (PFMT)/pelvic floor muscle exercise (PFME) on incontinence and birth trauma outcomes. Of these reviews, three were deemed to be of high quality (Haddow et al., 2005; Wagg & Bunn, 2007; Woodley et al., 2020), four of moderate quality (Khorasani et al., 2020; Lemos et al., 2008; Mantilla Toloza et al., 2024; Zhang et al., 2023) and one of low quality (Harvey et al., 2003) when critically appraised with the JBI Checklist for Systematic Reviews and Research Syntheses.

A systematic review of six RCTs examined prenatal and postnatal PFME on urinary incontinence and pelvic strength (Haddow et al., 2005). Participants in the included RCTs were females who had either a spontaneous or assisted vaginal delivery or non-elective caesarean. The results of this systematic review demonstrated positive findings for both prenatal and postnatal PFME for improving postpartum urinary incontinence. However, there was conflicting evidence of the clinical effectiveness of prenatal and postnatal PFME for preventing postpartum urinary incontinence (Haddow et al., 2005). In a separate systematic review, prenatal PFME was not found to prevent postnatal urinary incontinence at 3 months or improvements in pelvic floor strength (Harvey, 2003). There was also insufficient evidence to support the effectiveness of postnatal PFME in the prevention of faecal incontinence and pelvic floor prolapse (Harvey, 2003). In a systematic review and narrative synthesis of seven RCTs found that prenatal PFMT was effective in preventing urinary incontinence in the early postnatal period and improved pelvic floor muscle strength (Mantilla Toloza et al., 2024).

A Cochrane review published in 2020 of 46 trials (10,832 participants) assessed PFMT for the prevention of urinary incontinence and faecal incontinence in prenatal and postnatal women (Woodley et al., 2020). Prenatal PFMT was found to marginally decrease the risk of urinary incontinence during the mid-postpartum period (greater than 3-6 months postpartum) (29% less; RR 0.71, 95% CI 0.54 to 0.95; 5 trials, 673 participants; high-quality evidence).

Evidence was lacking to determine the impact of prenatal PFMT during the late-postpartum period (defined as greater than 6-12 months) (RR 1.20, 95% CI 0.65 to 2.21; 1 trial, 44 women; low quality evidence). Eight of the included trials assessed reported on faecal incontinence outcomes; however, only one trial assessed the prevention of faecal incontinence during the postpartum period. This found no evidence to support the effectiveness of postnatal PFMT in preventing faecal incontinence during the late postnatal period (RR 0.73, 95% CI 0.13 to 4.21; 1 trial, 107 women, low-quality evidence). Additional findings indicated no difference in the rates of caesarean sections, assisted deliveries or episiotomies between prenatal PFMT and control groups, and there was insufficient data to determine whether prenatal of postnatally delivered PFMT impacted quality of life. The systematic review demonstrated a positive association between prenatal and postnatal PFMT and pelvic floor muscle function (Woodley et al., 2020).

A systematic review and meta-analysis including four RCTs (n=675) assessing the clinical effectiveness of prenatal PFMT for the prevention of urinary incontinence reported a significant reduction in urinary incontinence from 6 weeks to 3 months post-delivery (odds ratio = 0.45; confidence interval: 0.3 to 0.66). The meta-analysis found no difference in urinary incontinence during the 34th and 35th gestational week of pregnancy (odds ratio = 0.13; confidence interval: 0.00 to 3.77) (Lemos et al., 2008). There was no significant difference in prenatal and postnatal perineal muscle strength following PFME (Lemos et al., 2008). Only one study included in the systematic review assessed PFME on health-related quality of life measured using the SF-36 and demonstrated a significant increase (p=0.004) in quality of life at 3 months postpartum (Lemos et al., 2008).

In a systematic review of four RCTs assessing the effectiveness of unassisted postnatal PFME for the prevention and treatment of urinary incontinence, one RCT included women who had a vaginal delivery, one RCT included a sample of women who had forceps or ventouse delivery, and the remaining two RCTs included women who had experienced both normal and assisted deliveries (Wagg & Bunn, 2007). Three of the included RCTs reported short-term improvements in already existing urinary incontinence, of which two RCTs demonstrated a statistically significant difference; however, this statistically significant difference was not reported at longer follow-up. The systematic review did not synthesise findings to report on the impact of postnatal PFMT for preventing incontinence. One RCT also reported improvements in pelvic floor muscle strength, but this finding was not statistically significant (Wagg & Bunn, 2007).

Another systematic review and meta-analysis assessed the effectiveness of prenatal PFMT alone or as part of a general physical activity program (Zhang et al., 2023). The analysis included 30 RCTs (6691 participants) and found that incorporating PFMT within exercise programs during pregnancy can prevent urinary incontinence (z = 3.46; p < 0.0005; relative risk [RR] = 0.72, 95% CI: 0.59, 0.87, I2 = 59%) and the incidence of third and fourth degree perineal tears ((z = 2.89; p = 0.004; RR = 0.50, 95% CI: 0.31, 0.80, I2 = 48%). The findings did not demonstrate a difference in episiotomy rates (Zhang et al., 2023).

A systematic review explored the clinical effectiveness of physiotherapy and pelvic floor muscle exercises on the prevention of incontinence among women who underwent natural childbirth or caesarean section (Khorasani et al., 2020). The systematic review included nine studies on the effects of prenatal physiotherapy and PFMT on the prevention of postpartum incontinence, of which six studies demonstrated positive findings on urinary incontinence prevention. One study included in the systematic review found that prenatal physiotherapy and PFMT was not effective in preventing faecal incontinence following childbirth.

Additionally, the systematic review did not demonstrate that physiotherapy and pelvic floor muscle exercises provide a reliable improvement in pelvic organ prolapse (Khorasani et al., 2020).

#### 2.2.2 Mixed exercise programmes with a pelvic floor muscle training element

Four systematic reviews (including one review of existing reviews) assessed the effectiveness of mixed exercise programmes with an included pelvic floor muscle training (PFMT) element. Of these reviews, two were deemed to be of high quality (Santos et al., 2024; Ryhtä et al., 2023) and two were of moderate quality (Perales et al., 2016; Davenport et al., 2018) when critically appraised with the JBI Checklist for Systematic Reviews and Research Syntheses.

In a review of existing reviews conducted by Ryhtä and colleagues, two included meta-analyses assessed the impact of exercise combined with PFMT during pregnancy for the prevention of urinary incontinence and reported a reduced risk of developing incontinence in the postpartum period (OR 0.63, 95% CI 0.51–0.79, I^2^=0%)(odds ratio (OR) 0.45, 95% CI 0.31–0.66, p < 0,0001, I^2^=7%) (Ryhtä et al., 2023). Prenatal exercise and PFMT was also found to reduce urinary incontinence at 3-6 months postpartum (RR 0.75, 95% CI 0.56–1.02, p=0.028). However, there was no evidence to show the impact of prenatal exercise and PFMT for the prevention of urinary incontinence in the long term (>5 years following childbirth) (RR 1.07, 95% CI 0.77–1.48, I^2^=25%) (Ryhtä et al., 2023).

Davenport et al (2018) conducted a systematic review and meta-analysis of 24 studies (15,982 participants) to assess the effectiveness of prenatal exercise (including but not limited to PFMT) on prenatal and postnatal urinary incontinence (Davenport et al., 2018). PFMT with or without aerobic training was found to reduce the risk of prenatal urinary incontinence (15 RCTs, 2764 participants; OR 0.50, 95% CI 0.37 to 0.68, I2=60%) and urinary incontinence during the postpartum period (10 RCTs, 682 participants; OR 0.63, 95% CI 0.51, 0.79, I2=0%), but the authors graded the quality of the evidence as low to moderate (Davenport et al., 2018).

A systematic review of 61 RCTs reported strong evidence (defined by the authors as ≥3 high-quality RCTs reporting on the outcome and ≥75% reporting a significant benefit) on the effectiveness of combined aerobic and resistance exercise during pregnancy on the prevention of urinary incontinence (Perales et al., 2016). Three RCTs reported a lower incidence of caesarean sections among trained women versus controls, but 12 RCTs did not find any significant differences (Perales et al., 2016). Another systematic review investigated the benefits of prenatal group aerobic and/or resistance training associated with PFMT (Santos et al., 2024). There was no difference in prevention of urinary incontinence between the exercise intervention and usual care, however the grade of this evidence was reported as low in the systematic review (RR: 0.57; 95% CI: 0.24−1.34; one study, 762 participants, random effects: p=0.20, subgroup for analysis at 3 months RR: 0.82; 95% CI: 0.56−1.19, one study, 722 participants, random effects: p = 0.30)(Santos et al., 2024).

#### 2.2.3 Prenatal perineal massage

Five systematic reviews assessed the effectiveness of prenatal perineal massage on incontinence and birth trauma outcomes. Of these reviews, one (a review of existing reviews) was deemed to be of high quality (Sananès et al., 2023) and three of moderate quality (Abdelhakim et al., 2020; Chen et al., 2022; Milka et al., 2023), and one was of low quality (Van Kampen et al., 2015) when critically appraised with the JBI Checklist for Systematic Reviews and Research Syntheses.

A meta-analysis including eleven RCTs (3467 participants) found no significant difference in postnatal urinary incontinence between the prenatal perineal massage group and control group (RR = 0.90, 95% CI [0.75, 1.09], p = 0.27), there was a significantly lower incidence of episiotomies (RR = 0.79, 95% CI [0.72, 0.87], p < 0.001) and perineal tears (RR = 0.79, 95% CI [0.67, 0.94], p = 0.007) (Abdelhakim et al., 2020).

A meta-analysis including 16 RCTs (6487 participants) found that there was no significant difference between prenatal perineal massage and the control group in incidence of urinary incontinence (RR = 0.91, 95% CI [0.79-1.05], P = 0:21) or faecal incontinence (RR = 0.75, 95% CI [0.51-1.11], P = 0:15) at 3 months postpartum (Chen et al., 2022). There was also no significant difference in the incidence of first- or second-degree perineal tears (RR = 0.96, 95% CI [0.90, 1.03], P = 0.30). The systematic review did find a significant difference in the incidence of third and fourth perineal tears between intervention and control groups (RR = 0.56, 95% CI [0.47, 0.67], p<0.00001) and postpartum pain at 3 months (RR = 0.64, 95% CI [0.51, 0.81], P = 0.0002) favouring the prenatal perineal massage group (Chen et al., 2022).

In a systematic review and narrative synthesis of 18 publications, the findings concluded that perineal massage reduces the incidence of perineal injuries and the risk of faecal and gas incontinence but did not reduce the risk of urinary incontinence (Milka et al., 2023).

A review of Cochrane systematic reviews of various prenatal, intrapartum and postpartum interventions for preventing postpartum urinary and faecal incontinence (Sananès et al., 2023), identified one meta-analysis that did not demonstrate a significant difference in either urinary incontinence (RR 0.90 95% CI 0.74 to 1.08) or faecal incontinence (RR 0.70 95% CI 0.27 to 1.80) following prenatal perineal massage (Beckmann, 2013).

In a systematic review exploring the effectiveness of different physiotherapy modalities during pregnancy, one included RCT reported no significant effect of prenatal perineal massage on the prevention of urinary, faecal or gas incontinence at 3-months postpartum (Van Kampen et al., 2015). Three separate studies included in the systematic review demonstrated a significant reduction in the incidence of second- or third-degree perineal tears and episiotomy rates following prenatal perineal massage; however, no differences in perineal pain were reported (Van Kampen et al., 2015).

#### 2.2.4 Pushing technique and vaginal device interventions

One systematic review of pushing technique and three systematic reviews of vaginal devices reported on incontinence, birth trauma outcomes and pelvic floor muscle strength. Of these systematic reviews, two were deemed to be of high quality (Oblasser et al., 2015; Shinozaki et al., 2022), one of moderate quality (Schreiner et al., 2018) and one of low quality (Harvey, 2003) when critically appraised with the JBI Checklist for Systematic Reviews and Research Syntheses.

A systematic review and meta-analysis of 17 RCTs (4606 participants) assessed the impact of pushing technique among primiparous women on urinary incontinence and birth trauma outcomes (Shinozaki et al., 2022). Results demonstrated a significant difference in urinary incontinence scores from baseline to postpartum in the spontaneous pushing group compared with the directed pushing group (two studies; 867 participants; standardised mean difference: –0.18). In terms of birth trauma outcomes, the systematic review found that spontaneous pushing led to a significant decrease in the need for suturing. However, there was no significant difference in the rates of severe perineal tears (third or fourth degree) or episiotomy rates between groups (Shinozaki et al., 2022).

Oblasser and colleagues systematically reviewed postnatal interventions using vaginal cones and balls to prevent incontinence in the postnatal period (Oblasser et al., 2015). Interventions for women in the postpartum period using cones and balls for any frequency or duration, combined with or without exercise, with any method of instruction (self-taught or instructions provided by healthcare practitioners) were eligible for inclusion. The systematic review aimed to identify evidence on the impact of vaginal cones and balls on urinary incontinence, pelvic floor muscle performance, and the following secondary outcomes: perineal descent or pelvic organ prolapse, adverse effects such as pain, and health economic outcomes including the cost of intervention time or teaching time. RCTs of women who had given birth by any method (vaginal, caesarean or assisted), regardless of whether they had any injuries during childbirth, were eligible for inclusion. This systematic review did not present evidence on the effectiveness of vaginal cones or devices to prevent urinary incontinence. Only one RCT (192 participants) was included in the review, which assessed the impact of interventions using vaginal cones to improve urinary incontinence (Oblasser et al., 2015). This review included a re-analysis of the raw data from the included RCT and found that the cone intervention demonstrated a significantly lower incidence of urinary incontinence at 12 months postpartum (RR 0.63, 95% CI 0.40–0.998 p=0.022), compared to the control group, but the results were limited due to the high degree of attrition and detection bias for the urinary incontinence outcome (Oblasser et al., 2015). In addition, a separate systematic review and narrative synthesis of PFME during and after pregnancy concluded that postnatal PFME when performed with a vaginal device with resistance or feedback can decrease postpartum urinary incontinence, but this systematic review did not report on the prevention of urinary incontinence (Harvey, 2003).

In a systematic review of pelvic floor interventions during pregnancy, three RCTs (1136 participants) studied the effectiveness of the EPI-NO perineal dilator (Schreiner et al., 2018). All three studies assessed different outcome measures, including perineal, anal sphincter and pelvic floor trauma, incidence of episiotomy and vaginal infection. Although incontinence was an included outcome in the systematic review, incontinence was not reported as an outcome measure in any of the included trials assessing the effectiveness of the EPI-NO perineal dilator. One RCT reported a significant difference in perineal trauma between the intervention and control group, favouring the EPI-NO group (intact perineum, 37.4% vs 25.7%; P=0.05). No significant differences were observed for any of the outcome measures reported (Schreiner et al., 2018).

#### 2.2.5 Bottom line evidence on the clinical effectiveness of pelvic floor muscle training and exercise, prenatal perineal massage, pushing technique and vaginal devices on incontinence

Twelve systematic reviews of pelvic floor muscle training (PFMT) and mixed exercise modes (with a PFMT element) were identified in this rapid review. Of the eleven systematic reviews assessing prenatal PFMT and exercise interventions, eight systematic reviews reported findings to support PFMT and mixed exercise for the prevention of urinary incontinence in the early postnatal period (up to 6 months postpartum). Evidence from two meta-analyses reported on longer-term incontinence and did not find evidence to support the effectiveness of prenatal PFMT and exercise to prevent urinary incontinence in the late postpartum period (defined as greater than 6-12 months) or after 5 years following childbirth. Two systematic reviews assessed the effectiveness of postnatal PFMT on urinary incontinence; however, one of the systematic reviews assessed effectiveness in terms of already existing incontinence but did not synthesise findings to report on the impact of postnatal PFMT for preventing incontinence. The other systematic review included a narrative synthesis of postnatal PFMT and reported conflicting evidence on its effectiveness to prevent urinary incontinence. Findings from three systematic reviews (two of postnatal PFMT and one of prenatal PFMT) found no evidence to support the effectiveness of PFMT to prevent postnatal faecal incontinence.

Five systematic reviews (including three systematic reviews and meta-analyses) explored the effectiveness of prenatal perineal massage for preventing incontinence. None of the meta-analyses found any significant differences in incidence of urinary incontinence (evidence from three meta-analyses) or faecal incontinence (evidence from two meta-analyses) following prenatal perineal massage.

Two systematic reviews reported on the effectiveness of vaginal devices for existing incontinence but did not report on the prevention of incontinence. In a systematic review of pushing technique, results demonstrated a significant difference in urinary incontinence scores from baseline to postpartum in the spontaneous pushing group compared with the directed pushing group.

### 2.3 Cost-effectiveness of interventions to prevent incontinence

All three of the included economic evaluations were deemed to be of high quality when appraised using the JBI checklist for economic evaluations (Joanna Briggs Institute, 2017a).

The first economic evaluation reports a within-trial cost-consequence analysis and cost-effectiveness analysis of a multi-centre randomised controlled trial to assess whether a policy of adopting an ‘upright position’ throughout second-stage labour increases the incidence of spontaneous vaginal delivery compared with a policy of adopting a ‘lying-down’ position among nulliparous women with epidural analgesia (Bick et al., 2017). Despite not being a directly relevant intervention to prevent incontinence, this economic evaluation of birthing positions collected short-term (3 months follow-up) and longer-term (1 year follow-up) outcomes of urinary and faecal incontinence following each delivery mode as part of the cost-consequence analysis. The analysis was undertaken from an NHS perspective over a one-year time horizon (Bick et al., 2017). Costs were health service utilisation costs presented in British pounds sterling for cost year 2013/14 (Table 3, Section 6.2).

The cost-effectiveness analysis of the delivery modes calculated the cost per additional case of spontaneous vaginal delivery where the lying down position was used as the comparator in the incremental cost-effectiveness ratio (ICER) calculation. The reported ICER (95% CI) was £722 (–£2968 to £6358) per additional case of spontaneous vaginal delivery for birthing in the upright position versus lying down (Bick et al., 2017). Incontinence was not considered as an outcome in this cost-effectiveness analysis. Therefore, priority was given to the presentation of the findings of the cost-consequence analysis as it reported on short and long-term incontinence outcomes following childbirth.

The trial demonstrated a statistically significant difference (adjusted risk ratio 0.86, 95% CI 0.78 to 0.94) in the rate of spontaneous vaginal deliveries between the groups with 41.1% of women who adopted a ‘lying down’ position achieving a spontaneous vaginal delivery, compared with 35.2% of women who remained in an upright position (Bick et al., 2017).

At 3 month follow-up, birthing position demonstrated no statistically significant difference on the prevalence of urinary incontinence (49.2% upright position, 49.4% lying down position; RR 0.99 (0.88 to 1.13)). However, a statistically significant difference in prevalence of faecal incontinence (defined as no bowel control and/or soiling) was observed at 3-months (11.5% upright position, 14.2% lying down position; RR 0.81 (0.59 to 1.12)). At 12 month follow-up, there was no statistically significant difference in urinary incontinence (as measured by the International Consultation on Incontinence Questionnaire-Urinary Incontinence) between birthing positions (Median [IQR]: 0 [0-4] upright position, 0 [0-4] lying down position; Median difference 0 [0 to 0]). No statistically significant difference was observed in prevalence of faecal incontinence between birthing position at this follow up (3.2% upright position, 3.2% lying down position; RR 1.02 (0.51 to 2.02)). Regarding immediate costs, the lying down position was associated with lower NHS resource use costs when giving birth at hospital, with a mean cost difference of £59 (95% CI £6 to £111) between groups. These lower resource use costs were due to the higher proportion of spontaneous vaginal deliveries in the lying down group compared with the upright position group. There was no significant difference in NHS resource use costs between groups for mothers and their infants at 12- months. Results of the cost-consequence analysis concluded that birthing position had little impact on the prevalence of urinary incontinence in the short-term, and no impact on prevalence in urinary or faecal incontinence at 12 months. A small statistically significant difference in prevalence of faecal incontinence was observed at 3 months.

The second economic evaluation was a cost-effectiveness analysis using data from a Cochrane systematic review of three different clinically effective models of pelvic floor muscle training (PFMT) for the prevention or treatment of postpartum incontinence (Brennan et al., 2021). Model 1 consisted of prenatal individually supervised PFMT to prevent urinary incontinence UI, model 2 was prenatal group-based PFMT to prevent or treat UI, and model 3 was postnatal individually supervised PFMT to treat urinary incontinence and prevent faecal incontinence. The economic evaluation was undertaken from the health service, consumer and societal perspectives and costs were presented in Australian dollars for cost year 2019 (Table 3, Section 6.2). The analysis presented the incremental cost of curing or preventing one case of incontinence. The incremental cost per case of urinary incontinence prevented or cured was $768 and $1,970 for model 1 and model 3, respectively. Model 2 (group-based PFMT) produced the greatest saving in costs (with $14.53 per case of urinary incontinence prevented or cured if eight women attended a session of group-based PFMT). For model 3, the cost to prevent or cure one case of faecal incontinence was $2,784 (Brennen et al., 2021). The authors concluded that group-based PFMT was found to be more cost-effective than individually supervised PFMT for treating women with postnatal incontinence (Brennen et al., 2021). However, it must be noted that the analysis did not compare cost-effectiveness of the intervention against a willingness to pay threshold making the results difficult to interpret by decision makers.

The third economic evaluation was a cost-utility analysis utilising a decision analytic Markov model of supervised prenatal PFMT and was conducted as part of the 2021 NICE guidance on the prevention and non-surgical management of pelvic floor dysfunction (NICE, 2021b). The proportion of women who would develop urinary incontinence over time in the absence of preventative PFMT was estimated using data from two studies reporting on the same intervention at different follow-up time-points, Reilly (2002), and Agur (2008), and the UR- CHOICE prediction model. The cost-utility analysis was conducted from an NHS and Personal Social Services perspective over a lifetime horizon. The cost of a PFMT session and the monthly health resource costs of urinary incontinence were presented in British Pounds Sterling for the cost year 2020. The cost-utility analysis found the intervention to be cost-effective compared to no intervention in a population with a 50% risk of pelvic floor dysfunction (NICE, 2021b). Total costs were greater in the group receiving the intervention (£827) than the group receiving no intervention (£539). The difference in costs was driven by the cost of administering the intervention. Urinary Incontinence management costs were similar across both groups, being slightly more costly in the group receiving no intervention (£539) than the group who received it (£507). The intervention group gained more QALYs (20.977) than the group not receiving the intervention (20.952). The corresponding incremental cost-effectiveness ratio (ICER) for the intervention group was £11,432 per QALY gained (NICE, 2021b). This indicates that the intervention is cost-effective at any cost-effectiveness threshold greater than £11,000. NICE typically recommends implementing interventions that have a cost per QALY between a threshold of £20,000 and £30,000.

#### 2.3.1 Bottom line evidence on the cost-effectiveness of pelvic floor muscle training and birthing position on incontinence

There is a dearth of evidence on the cost-effectiveness of interventions to prevent incontinence due to birth trauma. Evidence from one economic evaluation suggests that group-based pelvic floor muscle training (PFMT) during pregnancy is more cost-effective than postnatal individually supervised PFMT for preventing and curing urinary incontinence. However, the provision of PFMT for postnatal women experiencing urinary and faecal incontinence should not be discounted given its established effectiveness in preventing and treating both conditions. Evidence from a cost-utility analysis conducted by NICE utilising a decision analytic Markov model of supervised prenatal PFMT found the intervention to be cost-effective when compared to no intervention. The model utilised data from two randomised controlled trials and assumed a 50% risk of postnatal pelvic floor dysfunction in the population. The intervention was likely to be cost-effective for all willingness to pay thresholds over £11,000 per QALY gained. Although not a directly relevant intervention designed to prevent incontinence following childbirth, results of one economic evaluation of birthing positions concluded that birthing position had little impact on the prevalence of urinary incontinence at 3 months postnatal, and no impact on prevalence in urinary or faecal incontinence at 12 months. A small statistically significant difference in prevalence of faecal incontinence was observed at 3 months.

## 3. DISCUSSION

### 3.1 Summary of the findings

#### Clinical effectiveness

Incontinence is a debilitating and common condition faced by women following childbirth but can impact women at any stage of the life-course and is highly prevalent among middle-aged and postmenopausal women. NICE 2021 guidance (NG210) on the prevention and non-surgical management of pelvic floor dysfunction recommends pelvic floor muscle training (PFMT) for prenatal and postnatal women to prevent symptoms of incontinence which is one of the main symptoms of pelvic floor dysfunction (NICE, 2021a). The overwhelming majority of the evidence identified in this systematic review assessed the clinical effectiveness of PFMT and other exercise modes (with a PFMT element).

Of the eleven systematic reviews assessing prenatal PFMT and exercise interventions, nine systematic reviews reported findings to support PFMT and exercise for the prevention of urinary incontinence in the postnatal period (up to 6 months postpartum). Two of the included systematic reviews assessed the effectiveness of postnatal PFMT on incontinence; however, one of the systematic reviews assessed effectiveness in terms of already existing incontinence but did not synthesise findings to report on the impact of postnatal PFMT for preventing incontinence (Wagg & Bunn, 2007). The other systematic review included a narrative synthesis of postnatal PFMT and reported conflicting evidence on its effectiveness to prevent urinary incontinence (Haddow et al., 2005).

NICE advocates that early intervention in the postpartum period may decrease the risk of delayed-onset faecal incontinence in women; however, evidence is lacking on whether prenatal interventions (before obstetric trauma or injury) can provide a protective role in preventing faecal incontinence (NICE, 2007). Findings from three systematic reviews identified in this rapid review (one of prenatal PFMT and two of postnatal PFMT) found no evidence to support the effectiveness of PFMT to prevent postnatal faecal incontinence.

The 2021 NICE guidance indicates limited evidence supporting the long-term effectiveness of PFMT (NICE, 2021a). Our rapid review aligns with this assessment as two meta-analyses reporting on longer-term outcomes failed to demonstrate the effectiveness of PFMT and exercise to prevent urinary incontinence in the late postpartum period (defined as greater than 6-12 months; Woodley et al., 2020) or after 5 years following childbirth (Ryhtä et al., 2023).

Prenatal perineal massage was not discussed in the NICE 2021 guidance on the prevention and non-surgical management of pelvic floor dysfunction. Perineal massage during labour is recommended in the NICE 2023 guidance (CG190) on intrapartum care for healthy women and babies (NICE, 2023) as an intervention to prevent perineal trauma which is a known risk cause for continence issues (NICE, 2021a). This rapid review identified five systematic reviews (including three systematic reviews and meta-analyses) on the effectiveness of prenatal perineal massage for preventing incontinence. None of the meta-analyses found any significant differences in incidence of urinary incontinence (evidence from three meta-analyses) or faecal incontinence (evidence from two meta-analyses) following prenatal perineal massage.

The NICE 2021 guidance also notes that the effectiveness of intravaginal devices when combined with PFMT is still unclear based on current evidence and consequently the committee made a recommendation for research on the effectiveness of vaginal devices (NICE, 2021a). Our rapid review identified two systematic reviews reporting on the effectiveness of vaginal devices for existing incontinence, but neither of the reviews reported on the prevention of incontinence.

#### Cost-effectiveness

Incontinence is a devastating condition that places a significant financial burden on the healthcare system (Javanbakht et al., 2020). Costs from incontinence arise from various sources, including direct healthcare costs, patient out-of-pocket expenses for incontinence products and wider societal costs such as lost productivity due to absenteeism or reduced work performance (Fultz et al., 2005). The main focus of this rapid review was to identify evidence on the cost-effectiveness of interventions to prevent incontinence resulting from birth trauma. This rapid review identified three economic evaluation studies. Of which, two studies evaluated the cost-effectiveness of relevant interventions (PFMT) to prevent incontinence following childbirth (Brennen et al., 2021; NICE, 2021a). The third economic evaluation presented short and long-term postnatal incontinence outcomes but did not evaluate a relevant intervention designed to prevent incontinence (Bick, 2017).

Findings from a cost-effectiveness analysis suggest that group-based PFMT is more cost-effective for preventing urinary incontinence compared to individually supervised PFMT (Brennen et al., 2021). NICE conducted a cost-utility analysis as part of the 2021 NICE guidance on the prevention and non-surgical management of pelvic floor dysfunction (NICE, 2021b). The cost-utility analysis included a decision analytic Markov model to assess the cost-effectiveness of supervised prenatal PFMT compared with no intervention. The model utilised data from two randomised controlled trials and assumed a 50% risk of postnatal pelvic floor dysfunction in the population. The intervention was likely to be cost-effective for all willingness to pay thresholds over £11,000 per quality-adjusted life year (QALY) gained (NICE, 2021b).

NICE guidance states that prenatal supervised PFMT is likely to be cost-effective for some pregnant women, in particular women in the groups identified at higher risk of developing pelvic floor dysfunction (NICE, 2021a). The guidance recommends PFMT as an option as it is likely to be cost-effective for some women in these groups (NICE, 2021a).

### 3.2 Strengths and limitations of the available evidence

There is a dearth of evidence on the costs and cost-effectiveness of interventions to prevent incontinence resulting from birth trauma. Only three economic evaluations were identified in this rapid review; two of which assessed PFMT interventions (which were directly relevant to the rapid review question), and the other was an economic evaluation of birthing positions (an intervention not designed to prevent incontinence). The review failed to identify evidence on costs or cost-effectiveness for other types of interventions such as wider exercise programs, prenatal perineal massage, vaginal devices, or other intervention types.

There are challenges to evaluating the cost-effectiveness of interventions in this area, while QALYs are a valuable tool for measuring health-related quality of life in many contexts, their application to conditions like incontinence presents unique challenges. The impact of incontinence on health-related quality of life may vary across different population groups due to factors such as age and disability, as well as cultural and socioeconomic factors. In addition, due to the absence of a standard health outcome to incorporate within cost-effectiveness analyses of interventions to prevent incontinence, results of cost-effectiveness analyses in this area are difficult to interpret by decision makers due to the lack of a willingness to pay threshold.

There were a number of limitations in the included studies. Four of the systematic reviews were deemed to be of low quality when critically appraised using the JBI Checklist for Systematic Reviews and Research. Another limitation of the available evidence was a lack of information in the included systematic review papers on sample variables such as age, birthing delivery method and smoking status which restricted subgroup analyses in this review.

Clinical effectiveness was typically assessed over short term time horizons in most of the identified studies. The NICE manual for conducting health technology evaluations states a time horizon long enough to reflect all important differences in costs or outcomes should be used (National Institute for Health and Care Excellence, 2022). Given the potential for long-term impacts of incontinence, short time horizons may omit relevant costs or outcomes.

### 3.3 Strengths and limitations of this Rapid Review

This rapid review undertook thorough literature searches with no limit on search dates, using a well-developed search strategy and robust methodology. The searches aimed to identify evidence on the costs, cost-effectiveness and clinical effectiveness of any intervention type that aims to prevent incontinence resulting from childbirth. Despite the lack of available evidence on the cost-effectiveness of interventions to prevent incontinence due to birth trauma, this rapid review was successful in identifying a large evidence base on the clinical effectiveness of PFMT and exercise, although there was less evidence on prenatal perineal massage, birthing position and vaginal device interventions.

This review identified 20 systematic reviews assessing the clinical effectiveness of interventions. We acknowledge that by limiting the search for effectiveness evidence to systematic reviews only, there may be primary studies of additional intervention types to improve incontinence following childbirth that have not been represented in the identified systematic reviews. The systematic reviews included in this rapid review focused on specific intervention types such as PFMT, exercise and prenatal perineal massage. Only one of the included reviews evaluated ‘any type’ of intervention, this was a review of existing Cochrane systematic reviews of RCTs of any type of antenatal, intrapartum and postpartum interventions for preventing postpartum urinary and faecal incontinence (Sananès et al. 2023). However, there may be non-Cochrane systematic reviews of additional interventions that have not been captured in this review of reviews. In addition, there may also be primary studies of additional intervention types published since the date of the searches of the included reviews.

While reviews of existing reviews are valuable in synthesising evidence from multiple systematic reviews, including multiple reviews that include the same primary studies can lead to redundant information and inflated effect sizes. In addition, overlapping studies may differ in their methodological quality, which can increase the risk of bias. Different inclusion criteria across the different reviews make it difficult to compare and pool results. This rapid review utilised the Graphical Representation of Overlap for OVErviews (GROOVE) tool to assess the overlap of primary studies across the systematic review papers (Bracchiglione et al., 2022). The overlap was calculated at 3.5%, indicating ‘slight’ overlap across the studies (overlap cut-point grouping: <5% slight, 10% to <15% moderate, 10% to <15% high, >15% very high). Nevertheless, it must be acknowledged that the assessment of overlap conducted using the GROOVE tool only included 18 of the systematic reviews included in this rapid review. The two reviews of existing reviews were excluded from the assessment as they did not include a review of primary studies and therefore, it must be acknowledged that the degree of overlap of evidence may be greater than reported.

Due to the low number of economic studies in this topic area, the review was supplemented by targeted searches to present the economic case for investing in interventions that have been evidenced in terms of their clinical effectiveness. This involved a separate targeted search to identify the most useful studies reporting economic burden in this topic area (section 1.3). Nevertheless, we did not identify studies that directly assess the economic burden of incontinence directly related to perineal trauma in the UK.

This rapid review was limited to economic evaluations and systematic reviews of clinical effectiveness and therefore omits important qualitative evidence of women’s experiences and the acceptability of interventions to prevent incontinence. Consequently, a synthesis of qualitative evidence is needed in parallel to the evidence on the clinical effectiveness and cost effectiveness evidence in order to make recommendations for policy and practice.

### 3.4 Implications for policy and practice

Despite a paucity of economic evaluations assessing the cost-effectiveness of interventions for the prevention of incontinence, the substantial economic burden of incontinence on the NHS necessitates investment in clinically effective, preventative options. A large evidence base exists on the effectiveness of PFMT and exercise interventions to prevent incontinence due to childbirth, and the findings of this review present the case for investing in interventions that have been found to be clinically effective. This report synthesised evidence from twelve systematic reviews on the effectiveness of pelvic floor muscle training and exercise. However, the description, delivery and dosage of prescribed exercises were heterogenous across the reviews. While NICE emphasises the importance of ongoing pelvic floor muscle training for sustained benefits, low adherence rates may contribute to the limited evidence for long-term effectiveness. Consequently, assessments on the effectiveness of unsupervised PFMT and how best to support ongoing adherence such as the utilisation of e-health platforms are needed. It must also be noted that the provision of ongoing supervised pelvic floor muscle training will require additional resource allocation for staff time. Moreover, future recommendations for policy and practice must also consider qualitative findings of women’s experiences and the acceptability and feasibility of rolling out interventions for the prevention of incontinence which play an important role in the adherence and maintenance of PFMT.

### 3.5 Implications for future research

Regarding clinical effectiveness, PFMT and PFME were well represented in the evidence base identified in our rapid review. Other interventions, such as prenatal perineal massage and other therapeutic interventions, were less represented in this rapid review, especially for faecal incontinence outcomes, indicating a potential gap in the evidence base. Our rapid review findings are in-line with the recommendation for research on the effectiveness of vaginal devices made by NICE, our rapid review identified two systematic reviews reporting on the effectiveness of vaginal devices for existing incontinence, but neither of the reviews reported on the prevention of incontinence. We also acknowledge that by limiting the search for effectiveness evidence to systematic reviews only, there may be primary studies available assessing additional intervention types to improve incontinence following childbirth that have not been included in our rapid review.

A significant evidence gap exists regarding the cost-effectiveness of interventions aimed at preventing incontinence resulting from birth trauma. This rapid review indicates the necessity for economic evaluations to assess the value for money of these interventions. By quantifying the costs and benefits of different approaches, researchers can provide crucial information to policy makers to inform resource allocation decisions. Economic evaluations are required to assess the cost-effectiveness of interventions found to be clinically effective in the existing systematic reviews identified in this rapid review. The model-based cost-utility analysis undertaken by NICE is an example of this, utilising data from two previously published randomised controlled trials assessing the same intervention at different follow-up periods (NICE, 2021b).

Incontinence is a long-term burden as pregnancy and childbirth can weaken the pelvic floor, making women more susceptible to incontinence in later life (NICE, 2021a). Menopause often exacerbates these issues due to hormonal changes and by further weakening the pelvic floor muscles (Menezes et al., 2010). Obstetric trauma from vaginal delivery resulting in irreversible traumatic lesions to the urinary continence system is also a known link to incontinence in later life (Fritel et al., 2012). The evidence review conducted as part of the NICE 2021 guidance only identified one study on PFMT for women over 60 (not included in this rapid review as it did not meet the rapid review eligibility criteria) and made recommendations for future research in this area (NICE, 2021a). In their 2021 guidance, NICE also draws attention to the limited evidence on the long-term effectiveness of PFMT interventions (NICE, 2021a).

To fully understand the effectiveness and cost-effectiveness of interventions to prevent incontinence resulting from birth trauma, future research should adopt longer study time horizons to allow for the assessment of potential long-term impacts, such as the influence of interventions on incontinence during menopause. Exploration of outcomes over longer time horizons will enable researchers to gain valuable insights into any sustained benefits of interventions. Large scale studies of potentially inexpensive interventions are needed that better capture the wider societal costs and implications of the lifetime burden of incontinence. The use of model-based economic analyses can facilitate longer time horizons beyond that typically used in clinical trials through the extrapolation of cost and outcomes data. The model-based cost-utility analysis undertaken by NICE included a lifetime horizon in its analysis, achieved through simulating the data from two previously published randomised controlled trials (NICE, 2021b).

## Data Availability

All data produced in the present study are available upon reasonable request to the authors

## Abbreviations

| Acronym | Full Description |
| --- | --- |
| EQ-5D-3L | EuroQol-5 Dimensions, (three-level version) |
| FI | Faecal incontinence |
| CCA | Cost-consequence analysis |
| CI | Confidence interval |
| CINAHL | Cumulative Index to Nursing and Allied Health Literature |
| CRD | Centre for Reviews and Dissemination |
| CUA | Cost-utility analysis |
| GI | Gas incontinence |
| GROOVE | Graphical Representation of Overlap for OVERviews |
| ICS | International Continence Society |
| ICER | Incremental Cost-effectiveness ratio |
| IIQ-7 | Incontinence Impact Questionnaire Short Form |
| JBI | Joanna Briggs Institute |
| LOS | Length of Stay |
| NHS | National Health Service |
| NHS EED | The NHS Economic Evaluation Database |
| NICE | National Institute for Health and Care Excellence |
| OECD | Organisation for Economic Co-operation and Development |
| PFM | Pelvic floor muscles |
| PFEs | Peripartum pelvic floor exercises |
| PFMT | Pelvic floor muscle training |
| PFME | Pelvic floor muscle exercises |
| PA | Physical Activity |
| POP | Pelvic Organ Prolapse |
| PRISMA | Preferred Reporting Items for Systematic reviews and Meta-Analyses |
| QALY | Quality Adjusted Life Year |
| QoL | Quality of Life |
| RCT | Randomised controlled trial |
| REEDA | Redness, oedema, ecchymosis, discharge and anal incontinence scale |
| RR | Risk ratio |
| SF-12 | 12-Item Short Form Health Survey |
| SMD | Standard Mean Difference |
| SUI | Stress urinary incontinence |
| SVB | Spontaneous vaginal birth |
| UI | Urinary incontinence |
| UDI-6 | Urogenital Distress Inventory, Short Form |
| VAS | Visual Analog Scale |

## Glossary of key terms

**Prenatal:** a term that means ’before birth’ (’antepartum’ is an alternative term).

**Postnatal:** a term that means “after birth” and in this report describes the year following the birth of a baby (‘postpartum’ is an alternative term).

**Perinatal:** a term that means the period of time when a person becomes pregnant and up to a year after giving birth.

**Pelvic floor muscle training:** exercises that strengthen the muscles that support the bladder, rectum, and uterus.

**Primiparous:** a term used to describe a woman who has given birth once or who is experiencing her first pregnancy.

**Nulliparous:** a term used to describe a woman who has not given birth previously.

*inflated using Bank of England Inflation Calculator: https://www.bankofengland.co.uk/monetary-policy/inflation/inflation-calculator

## 5. RAPID REVIEW METHODS

### 5.1 Eligibility criteria

The eligibility criteria are described in Table 1.

**Table 1.** Eligibility criteria.

|  | **Inclusion criteria** | **Exclusion criteria** |
| --- | --- | --- |
| **Participants** | Prenatal and postpartum women | Women who are not prenatal and postpartum<br>Men, children |
| **Intervention / exposure** | Any intervention (pre- intra or post-natal) designed to prevent continence issues resulting from perineal birth trauma.<br>Interventions will include: Birthing position, perineal massage, digital perineal massage (performed by the woman or her partner), myofascial therapy, strengthening exercises (including Kegels), bladder training, nerve modulation, pelvic floor therapy, electrical simulation to improve pelvic floor muscle contraction, pessary, hyaluronidase, acupuncture. | Interventions promoting healing or treating perineal trauma not resulting from perineal birth trauma.<br>Surgical and medical interventions that are unsuitable or unfeasible to roll out e.g., planned caesareans, routine episiotomies.<br>Papers that do not report on an intervention such as screening strategies.<br>Papers that do not explicitly state that the intervention aims to prevent incontinence (urinary, faecal or gas incontinence). |
| **Comparison** | Usual care or any alternative intervention including placebo or no intervention. |  |
| **Outcomes** | Primary outcome: Incidence of continence issues<br>Any intermediate outcomes will be considered which may include, but are not limited to: birth trauma outcomes (e.g., perineal tear), quality of life, ability to continue to work, morbidity, mobility, perineal strength, pelvic floor strength, sexual function. | Incidence of post-partum infections or other risk factors.<br>Continence issues not relating to pregnancy or as a result of perineal birth trauma. |
|  | NB. Papers must explicitly state that the intervention aims to prevent continence issues even if the study does not report on continence issues as an outcome measure.<br>Cost-effectiveness outcomes will include: cost per incidence of incontinence prevented, cost per QALY gained, cost per disability-life year (DALY) averted. |  |
| **Study design** | Cost-effectiveness: Trial-based and model-based full economic evaluations (cost-effectiveness analyses, cost-utility analyses, cost-benefit analyses, cost-minimisation analyses). Partial economic evaluations (cost-consequence analyses, cost-analyses, cost-description studies, cost-outcome descriptions).<br>Clinical effectiveness: Systematic reviews and reviews of existing reviews/ umbrella reviews | Clinical effectiveness: Scoping reviews, rapid reviews, primary clinical effectiveness studies |
| **Countries** | A focus on UK and European literature with a view of broadening to OECD countries. | Non-OECD countries |
| **Language of publication** | English | Full text publications not available in the English language |
| **Publication date** | From inception |  |
| **Publication type** | Published and preprint |  |

### 5.2 Literature search

The search strategy conducted in Medline via Ovid is presented in Appendix 1. Dates of the searches were from inception until 7^th^ June 2024. Three search filters were incorporated within the search strategy: the CRD NHS EED filter for economic evaluation studies, the NICE OECD countries search filter (Ayiku et al., 2021), and a filter for systematic review studies. Searches were conducted in the following databases: Medline (Ovid), EMBASE (Ovid), CINAHL (EBSCO), EMCARE (Ovid), Cochrane Library, and CRD.

A separate targeted search was conducted to identify relevant studies on the economic burden of disease, which is presented as a separate section in the background section of this rapid review report to present the economic argument for investing in clinically effective interventions to reduce incontinence following pregnancy and perineal trauma. This search used the search strands from the main rapid review search strategy (Appendix 1) for the incontinence and pregnancy terms and the search filters for cost of illness studies for Medline and Embase presented in Chapter 20 of the Cochrane Handbook for Systematic Reviews of Interventions (Aluko et al., 2023). In addition to the searches conducted in Medline and Embase, supplementary searches were conducted in Google Scholar. As per the Cochrane guidance (Chapter 20, section 20.2.3), the evidence of economic burden is presented in the background section of this rapid review report and includes the few most useful articles that report information on the economic burden of the condition being addressed (Aluko et al., 2023).

### 5.3 Study selection process

Two reviewers independently screened 100% of the titles and abstract using the Covidence review management software. The same two reviewers then independently screened 100% of the full text articles. Following the independent full text screening stage, discrepancies were resolved through discussion between the two reviewers and the review lead (BFA) to come to an agreement on the final inclusions if there was ongoing disagreement.

### 5.4 Data extraction

Data extraction was based on the outlined eligibility criteria. For the economic evaluation studies, the review team extracted data on study country, type of intervention, birthing delivery method, study design, sample size, length of follow-up, type of economic evaluation, perspective of analysis, currency and cost year, details of discounting and sensitivity analysis, main costs and outcomes measures, and main health economics findings.

For the systematic review studies, the following data extraction variables were included: lead author institution, review period, review aim, included study designs, included outcome measures, number of included studies, characteristics of included studies (study designs, countries, intervention type, birthing delivery method), and key findings. All four members of the core BIHMR/CHEME review team completed the data extraction, and the review lead (BFA) conducted the final verification of the data extraction.

### 5.5 Quality appraisal

Economic evaluation studies were assessed using the JBI critical appraisal checklist for economic evaluations, and the systematic reviews were assessed using the JBI Checklist for Systematic Reviews and Research Syntheses (Joanna Briggs Institute, 2017b).

### 5.6 Synthesis

Due to the heterogeneity of the included studies, a narrative synthesis was reported.

## 6. EVIDENCE

### 6.1 Search results and study selection

After the removal of duplicates, the search identified 3,383 studies. Full texts (n=56) were reviewed, and 23 studies were included in this rapid review: systematic reviews (n=20), economic evaluations (n=3).

### 6.2 Data extraction

Data extraction for the systematic reviews and the economic evaluations are presented in Table 2 and Table 3, respectively.

**Table 2:** Evidence from included systematic reviews of clinical effectiveness (n=20).

| Citation and country of lead author institution | Review details | Key characteristics of included studies | Quality | Key findings |
| --- | --- | --- | --- | --- |
| Abdelhakim et al., 2020<br>Egypt | **Review period:** Inception to August 2019<br>**Review purpose:** This systematic review and meta-analysis aimed to update the current evidence base about whether antenatal perineal massage reduces the risk of perineal trauma and postpartum complications.<br>**Included study designs:**<br>Randomised Controlled Trials (RCT)<br>**Included outcome measures:**<br>Primary outcome measure was the risk of all degrees of perineal tears and the incidence of episiotomies. Secondary outcome measures include the duration of second stage of labour in hours, perineal pain as evaluated by VAS, wound healing as evaluated by redness, edema, ecchymosis, discharge and anal incontinence (REEDA) scale, urinary incontinence and anal incontinence (faecal and flatus incontinence) reported within 3 | **Number of included studies: 11**<br>**Key characteristics:**<br>**Study design:** All n=11 included studies were RCTs that compared perineal massage versus no perineal massage during antenatal care. All studies performed antenatal digital perineal massage in last 4 to 6 weeks before delivery. Massages conducted either by pregnant women or partners.<br>**Countries:** Egypt (n=4), Canada (n=2), Nigeria (n=1), Turkey (n=1), Iran (n=1), Japan (n=1), UK (n=1).<br>**Intervention type:** Digital Antenatal perineal massage (n=11).<br>**Delivery method:** Vaginal birth (n=11) | **Quality Rating - Moderate (6/11)** | Antenatal perineal massage significantly reduced the incidence of episiotomies and perineal tears, especially third and fourth-degree perineal tears.<br>Prenatal perineal massage caused a significant decrease in the second stage of labour duration, postpartum perineal pain, and anal incontinence. No significant difference was identified in urinary incontinence between usual care and antenatal perineal massage. |
|  | months postpartum, and Apgar scores at 1 and 5 minutes. |  |  |  |
| Chen et al., 2022<br>China | **Review period:** Inception to April 2022<br>**Review purpose:** This systematic review and meta-analysis sought to update the evidence base on whether prenatal perineal massage can reduce the risk of perineal trauma and postpartum complications.<br>**Included study designs:** RCTs.<br>**Included outcome measures:** Primary measures were the risk of perineal tear, incidence of perineal incision and vaginal delivery. Secondary outcome measures were perineal pain (assessed by VAS), urinary and faecal incontinence at 3 months postpartum. | **Number of included studies:** 16<br>**Key characteristics:**<br>**Study design:** All 16 studies were RCTs comparing perineal massage versus no perineal massage during prenatal care.<br>**Countries:** Egypt (n=4), Canada (n=2), Austria (n=1), Turkey (n=1), Australia (n=1), United States (n=1), UK (n=1), Spain (n=1), Ireland (n=1), Iran (n=1), Japan (N=1) and Nigeria (n=1).<br>**Intervention type:** Prenatal perineal massage.<br>**Delivery method:** Vaginal delivery (n=16). | **Quality Rating – Moderate (6/11)** | Prenatal perineal massage was found to significantly reduce the incidence of perineal tears and episiotomy, especially for third and fourth-degree perineal tears. Furthermore, prenatal perineal massage may significantly reduce the incidence of perineal pain three months after delivery.<br>No significant difference in terms of incidence of vaginal delivery, perineal pain, urinary incontinence and faecal incontinence between prenatal perineal massage intervention group and control group (no prenatal perineal massage).<br>Prenatal perineal massage could reduce the risk of perineal incision.<br>Obstetrics and gynaecology professionals should consider popularizing prenatal perineal massage. |
| Davenport et al., 2018<br>Canada | **Review period:** Inception – 6 January 2023<br>**Review purpose:** To examine the relationships between prenatal physical activity and prenatal and postnatal urinary incontinence (UI).<br>**Included study designs:** Primary studies of any design were eligible, with the exception of case studies (n=1), narrative syntheses and systematic reviews<br>**Included outcome measures:** Prevalence and symptoms of UI during the prenatal and/or postpartum period (up to 12 months postpartum). | **Number of included studies:** 24<br>**Study design:** Study designs of included articles were Randomised Controlled Trials (n =18), non-randomised intervention (n= 2) and cohort studies (n =4).<br>**Countries:** Not stated in paper and supplementary files unavailable. Paper reports that studies were conducted in 12 countries and 4 continents.<br>**Intervention type:** Prenatal exercise of any frequency, intensity, duration, volume, or type.<br>**Delivery method:** Not reported, unable to access supplementary files. | **Quality Rating – Moderate (8/11)** | Prenatal pelvic floor muscle training (PFMT) with or without aerobic exercise decreased the odds of UI in pregnancy (15 randomised controlled trials (RCTs), n=2764 women; OR 0.50, 95% CI 0.37 to 0.68, I<sup>2</sup> =60%) and in the postpartum period (10 RCTs, n=1682 women; OR 0.63, 95%CI 0.51, 0.79, I<sup>2</sup> =0%). Exercise was beneficial at preventing the development of UI in women with continence, but not effective in treating UI in women with incontinence. Prenatal exercise had a moderate effect in the reduction of UI symptom severity during (five RCTs, standard mean difference (SMD) -0.54, 95%CI -0.88 to -0.20, I<sup>2</sup> =64%) and following pregnancy (three RCTs, 'moderate' quality evidence; SMD -0.54, 95% CI -0.87 to -0.22, I<sup>2</sup> =24%). |
| Haddow et al. 2005<br>Australia | **Review period:** 1981 to 2003<br>**Review purpose:** To determine, from the available evidence, the effectiveness of an antenatal and/or a post-natal program of pelvic floor muscle exercises (PFME) compared with usual care on preventing, reducing or resolving the incidence and severity of stress incontinence, urge incontinence or mixed stress and urge urinary incontinence following childbirth. Secondary objectives were included to examine the effectiveness of a PFME program on pelvic floor muscle strength and on encouraging adherence to an exercising program.<br>**Included study designs:** Randomised controlled trials and non-randomised controlled trials.<br>**Included outcome measures:** Outcomes that were of interest: non-occurrence of urinary incontinence following childbirth; a change in the frequency, duration or severity (as appropriate) of urinary incontinence up to 12 months following childbirth; a change in the strength of pelvic floor muscle contractions; period of time PFME continued after initial instruction; frequency of PFME undertaken; women's awareness of the importance of PFME; satisfaction with PFME instruction. | **Number of included studies:** 10<br>**Study design:** Study designs of included articles were randomised controlled trials (N=7), non-randomised controlled trials (N=3).<br>**Countries:** Australia (n=1), New Zealand (n=1), Norway (n=3), UK (n=1), USA (n=3), UK & New Zealand (n=1).<br>**Intervention type:** Pelvic floor muscle exercises: Antenatal PFME (n=5); postnatal PFME (n=5).<br>**Delivery method:** Postnatal PMFE (n=5): normal vaginal birth (n=4), Forceps or ventouse deliveries (n=1) | **Quality Rating – High (10/11)** | In terms of the effectiveness of PFME programs, the results of this review indicate that urinary incontinence following childbirth can be improved by performing PFME and that any form of a specific PFME program appears to improve exercising frequency. However, the value of individual components of PFME programs, such as take-home materials, reminder telephone calls and feedback of exercising effectiveness, is less clear.<br>It is reasonable to conclude that contact with the health professionals providing the instruction and advice, either in the form of weekly telephone calls or in the form of monthly group sessions, affects the frequency with which women performed PFME and consequently the better urinary continence outcomes.<br>Although the review did not identify the specific number of occasions a PFME program needs to be provided to have a significant effect on urinary incontinence, at least two instruction sessions are suggested by the findings. |
| Harvey, 2003 | **Review period:** From inception to 2002. | **Number of included studies:** 9 | **Quality Rating – Low (5/11)** | Postpartum PFEs appear to be effective in decreasing postpartum urinary incontinence. |
| Canada | **Review purpose:** To review the literature on the origin, anatomical rationale, techniques, and evidence-based effectiveness of peripartum pelvic floor exercises (PFEs) in the prevention of pelvic floor problems including urinary and anal incontinence and prolapse.<br>**Included study designs:** Randomised controlled trials.<br>**Included outcome measures:** Presence of postpartum urinary incontinence as determined by self-report, urodynamic studies, standardized pad test, or diary; pelvic floor strength as evaluated by perineometry; and self-report of anal (flatal and faecal) incontinence. | **Study design:** Study designs of included articles were randomised controlled trials (n=9).<br>**Countries:** Canada (n=1), Norway (n=1), UK (n=2), USA (n=2), Australia (n=1), Sweden (n=1), UK & New Zealand (n=1).<br>**Intervention type:** Antenatal PFE (urinary incontinence) (n=3); postnatal PFE (urinary incontinence) (n=4). Postnatal PFE (anal incontinence) (n=2)<br>**Delivery method:** N/A for antenatal studies (n=3); normal vaginal birth (n=5); assisted delivery with instruments (n=1) |  | Data regarding the effect of PFEs on prevention of anal incontinence are lacking, and also on its prevention of prolapse.<br>The studies published had inherent limitations that impacted on the application of PFEs antepartum and postpartum. None of the studies had limited entry criteria to women continent at baseline. Outcome measures were not standardised, in particular with the subjective self-report of urinary incontinence. Very little data were available regarding the preventive role of PFE in faecal incontinence, whereas data were totally absent with respect to pelvic organ prolapse. |
| Khorasani et al., 2020<br>Iran | **Review period:** Inception until December 2017<br>**Review purpose:** This review aimed to examine the effects of physiotherapy and PFME compared to no interventions in preventing and treating pregnancy-related pelvic floor dysfunctions.<br>**Included study designs:** RCTs or studies with quasi-RCT design.<br>**Included outcome measures:** prevalence and incidence of pelvic organ prolapse (POP), urinary incontinence (UI) and faecal incontinence (FI) 3 to 12 months postpartum. | **Number of included studies: 26**<br>**Key characteristics:**<br>**Study design:** n=26 RCTs, n=25 were 2-arm RCTs (intervention and control groups) and n=1 was a 3-arm RCT (1 single intervention, 1 combined intervention and 1 control group).<br>**Countries:** UK (n=5), New Zealand (n=1), Australia (n=1), Norway (n=3), Ireland (n=2), The Netherlands (n=1), Turkey (n=3), South Korea (n=2), Brazil (n=1), Spain (n=1), Sweden (n=1), Canada (n=1), France (n=1), Thailand (n=1), USA (n=1) and China (n=1).<br>**Intervention type:** All n=26 included studies reported pelvic floor muscle training/exercise interventions. (n=3) reported Kegels and (n=4) reported the<br>sessions were conducted with a physiotherapist.<br>**Delivery method:** (n=13) vaginal; (n=6) prepartum, unclear /not reported (n=7) | **Quality Rating – Moderate (8/11)** | UI severity and prevalence was reported in nine studies. UI prevalence in PFME intervention groups had significantly improved in six included studies. No significant difference was observed between groups in terms of UI prevalence in the remaining (n=3) studies.<br>N=1 included study reported individuals receiving the PFME intervention reported lower prevalence of FI than those in the control arm, however this difference was not significant. |
| Lemos et al., 2008<br>Brazil | **Review period:** 1966 to 2007<br>**Review purpose:** The aim of the current article was to conduct a systematic review of the performance of perineal exercises during pregnancy and their utility in the prevention of urinary incontinence.<br>**Included study designs:**<br>Randomised Controlled Trials<br>**Included outcome measures:**<br>Pelvic floor strength, type, frequency, intensity and diligence of the prescribed exercises, quality of life measures. | **Number of included studies:** 4<br>**Key characteristics**<br>**Study design:** Randomised Controlled Trials (n = 4)<br>**Countries:** US (n = 1), UK (n = 1), Norway (n = 1), Mexico (n = 1)<br>**Intervention type:** Isolated perineal exercise programs, without the use of any other kind of device, during pregnancy for the prevention of urinary incontinence.<br>**Delivery method:** N/A – (prenatal n=4) | **Quality Rating – Moderate (8/11)** | Four RCTs with high methodological quality, involving a total of 675 women were included. They indicated that perineal muscle exercise significantly reduced the development of urinary incontinence from 6 weeks to 3 months after delivery (odds ratio = 0.45; confidence interval: 0.3 to 0.66). However, when evaluating this effect during the 34th and 35th gestational week, a meta-analysis showed that the results were not significant (odds ratio = 0.13; confidence interval: 0.00 to 3.77). |
| Mantilla Toloza et al., 2024<br>Columbia | **Review period:** Not reported in paper.<br>**Review purpose:** To determine the effectiveness of pelvic floor muscle training (PFMT) in the prevention of SUI in women during the antenatal and postnatal period by reviewing and evaluating the available scientific literature<br>**Included study designs:**<br>Randomised controlled trials.<br>**Included outcome measures:**<br>Eliminated SUI, asymptomatic dysfunction detected, interrupted progression of incontinence. | **Number of included studies:** 7<br>**Study design:** Study designs of included articles were randomised controlled trials (n=7).<br>**Countries:** UK (n=2), Norway (n=2), Poland (n=1), France (n=1), Brazil (n=1),<br>**Intervention type:** Antenatal PFME (n=6); postnatal PFME (n=1).<br>**Delivery method:** N/A for antenatal studies (n=6); no data reported for delivery method of the postnatal study and unable to access full text (n=1). | **Quality Rating – Moderate (7/11)** | The application of PFMT early in pregnancy has positive effects on urinary continence after childbirth. The application of protocols that include individualised instruction and adequate follow-up by a physical therapist allows women to increase adherence to pelvic floor training, as well as possibly promoting motivation and awareness for proper performance of these exercises during interventions and at home. A training protocol that follows the general principles of strength training, emphasising near-maximal contractions and at least a 6-week training period, emphasising strenuous intensity training, may be as effective as a 4month training at moderate intensity. It is important to develop new studies to determine the intensity, time of application, and number of<br>repetitions of each exercise for the prevention of SUI in late pregnancy and postpartum |
| Milka et al. 2023<br>Lead author institution: Poland | **Review period:** Inception – June 2023<br>**Review purpose:** The aim of the review was to assess and analyze the impact of APM (antenatal perineal massage) on perinatal perineal injuries and the development of pelvic pain and other complications in postpartum women, such as dyspareunia, urinary (UI), gas (GI), and faecal incontinence (FI).<br>**Included study designs:** There was no restriction on the study design.<br>**Included outcome measures:** Assessment of perinatal perineal injuries after APM - by medical personnel. Perineal pain - immediately after childbirth, during the postpartum period, VAS, VRS scale, verbal scale, e.g., no pain, medium, high, unbearable. Assessment of urinary/gas/faecal continence problems - proprietary questionnaire, standardized scales, e.g., KHQ, manometers, sonographic. Sexual dysfunctions - postpartum period, VAS scale, original questionnaires, ICIQ scales. | **Number of included studies: 18**<br>**Key characteristics:**<br>**Study design:** Study designs of included articles were non-randomised controlled designs (N=1), controlled clinical trials (N=2), RCT (N=13), observational studies (N=1), prospective controlled trial (N=1).<br>**Countries:** Spain (n=4), Austria (n=1), Brazil (n=3), Egypt (n=1), Ireland (n=1), Turkey (n=1), Canada (n=3), Israel (n=1), UK (n=1), Japan (n=1) and Nigeria (n=1).<br>**Intervention type:** Antenatal perineal massage (n=18).<br>**Delivery method:** Vaginal birth (n=18) | **Quality Rating – Moderate (7/11)** | APM performed in the second half of the third trimester of pregnancy is conducive to protecting the perineum during labor. Perineal massage during pregnancy reduces the risk of GI and FI in the puerperium. Unfortunately, a similar effect has not been demonstrated for UI. There are no unequivocal reports on the impact of APM on sexual dysfunction.<br>Techniques of APM should be constantly improved. Current information on performing APM are insufficient. There are no recommendations that say unequivocally about the best time to start a massage, its duration and frequency. Some researchers recommended only internal vaginal massage, some also external. All these factors affect the effectiveness of massage, which should be considered when designing further research on its impact on the state of a woman during labor and the postpartum period. |
| Oblasser et al., 2015<br>England, UK | **Review period:** Inception – September 2014<br>**Review purpose:** The objectives of this review were: to compare the effectiveness of vaginal cones or | **Number of included studies: 1**<br>**Study design:** RCT (n=1)<br>**Countries:** New Zealand | **Quality Rating – High (10/11)** | Authors of the one included study identified in the systematic review provided the raw data to the authors of the systematic review to conduct secondary analysis which comprise the findings presented in this review paper; however, health economics outcomes were not reported. |
|  | balls for improvement of pelvic floor muscle performance and urinary continence in the postpartum period to no treatment, placebo, sham treatment or active controls; to gather information on effect on perineal descent or pelvic organ prolapse, adverse effects and economical aspects.<br>**Included study designs:**<br>Randomised and quasi-randomised controlled trials with individual or cluster randomisation and parallel design.<br>**Included outcome measures:**<br>Primary outcomes: pelvic floor muscle performance (e.g. strength, endurance), determined using a valid and reliable measure, e.g. vaginal squeeze pressure or participant reported improvement; urinary(in)continence, determined using a valid and reliable measure, e.g. quantified symptoms or urodynamics.<br>Secondary outcomes: perineal descent or pelvic organ prolapse. Health economics, e.g., cost of interventions or teaching time, as determined in each of the included studies. | **Intervention type:** Enforced exercise regimen with physiotherapist with one training session and three follow-up visits and three, six, and 9-months postpartum: factorial design with three subgroups: pelvic floor muscle exercises, vaginal cones, and both.<br>**Delivery method:** All birth delivery modes were eligible for inclusion; however, data for the one included study was not reported in the systematic review, and full text of included study not available. |  | Results of the secondary analysis found that compared to the control group, the cone group shows a statistically significant lower rate of the primary outcome urinary incontinence at 12 months post-partum (RR 0.63, $p=0.022$), but an almost same rate of urinary incontinence in the cone group cannot be excluded (95% CI 0.40–0.998). Exploratory analyses of pad test and perineometry measurements do not support the difference found for urinary incontinence (all $p$-values$>0.05$). Compared to the exercise group, the prevalence of urinary incontinence in the cone group is similar (RR 1.01, $p=1.000$), but a prevalence of urinary incontinence half or almost twice as high in the cone group cannot be excluded (95% CI 0.52–1.93). Exploratory analyses of pad test and perineometry measurements support these findings (all $p$-values$>0.05$ showing no statistically significant difference between cone and exercise group). |
| Perales et al, 2016<br>Spain | **Review period:** Until May 2015<br>**Review purpose:** To understand what evidence exists with regard to maternal and offspring benefits of aerobic and/or resistance training during pregnancy. | **Number of included studies:** 61<br>**Key characteristics:**<br>**Study design:** Randomised Controlled Trials (n = 61)<br>**Countries:** Not stated | **Quality Rating – Moderate (7/11)** | **Aerobic + resistance exercise** Strong evidence for cardiorespiratory fitness with exercise interventions lasting 12 to 24 weeks. One RCT found an improvement in maternal muscle strength after only 6 weeks which lasted up to 6 weeks postpartum. Strong evidence on the benefits of combined exercise interventions for preventing urinary incontinence, with three of four studies reporting positive effect. Of note, the |
|  | **Included study designs:**<br>Randomised Controlled Trials<br>**Included outcome measures:**<br>Impact of aerobic exercise on, maternal gestational weight gain, cardiorespiratory fitness, preventing urinary incontinence, maternal muscle strength, rates of caesarean sections, duration of the first labour stage, reduction in sick leave, risk of hypertension. | **Intervention type:** Aerobic (n = 15), resistance (n = 6) or combined (aerobic + resistance) exercise interventions (n = 32), or exercise counselling interventions (n = 8).<br>**Delivery method:** NA – all studies were antenatal. |  | aforementioned RCTs also included specific pelvic floor muscle training. |
| Ryhtä et al, 2023<br>Finland | **Review period:** Inception to 11 January 2023.<br>**Review purpose:** To summarise the existing evidence about the effectiveness of exercise interventions on urinary incontinence and pelvic organ prolapse (POP) in pregnant and postpartum women.<br>**Included study designs:**<br>Systematic reviews and/or meta-analysis.<br>**Included outcome measures:**<br>Incidence of UI, prevalence and severity of UI, incontinence, prolapse, pelvic floor strength, prevalence of UI, FI, incontinence specific QoL, POP symptoms. | **Number of included studies:** 9 systematic reviews, reporting findings from 89 original studies.<br>**Key characteristics:**<br>Study design: Systematic reviews (n = 9)<br>**Countries:** Switzerland (n = 1), Canada (n = 3), Brazil (n = 1), Norway (n = 1), UK (n = 2), New Zealand (n = 1)<br>**Intervention type:** Any type of physical activity (e.g., exercise, physiotherapy) or guidance to address the symptoms of UI and/or POP, delivered individually, in a group, face-to-face, or online.<br>**Delivery method:** Not stated. | **Quality Rating – High (11/11)** | The highest level of evidence was found for preventing the symptoms of postpartum urinary incontinence through exercise and pelvic floor muscle training pelvic floor muscles (PFMT) during pregnancy. Moderate-level evidence showed that exercise and PFMT are likely to reduce the symptoms and severity of urinary incontinence, but the level of evidence was low on PFMT reducing the symptoms of POP.<br>Encouraging and guiding pregnant and postpartum women to exercise and train PFM and identifying pregnant and postpartum women with symptoms of PFM dysfunction and directing them to a physiotherapist or other health care professional specializing in pelvic floor function is recommended. |
| Schreiner et al., 2018<br>Brazil | **Review period:** January 1990 to December 2016<br>**Review purpose:** to assess published randomised trials on pelvic floor interventions during pregnancy in healthy women and evaluate the consequences for | **Number of included studies:** 22<br>**Key characteristics:**<br>**Study design:** Study designs of included articles were randomised controlled trials (n=22). | **Quality Rating – Moderate (7/11)** | The use of the EPI-NO device was not superior to the usual recommended prenatal follow-up for reducing pelvic floor lesions. However, several clinical trials showed evidence that PFMT shortens the second stage of labour and reduces the risk of urinary incontinence during pregnancy. Therefore, PFMT should be taught routinely during prenatal care and should be |
|  | childbirth and pelvic floor dysfunctions.<br>**Included study designs:**<br>Randomised controlled trials with healthy pregnant women.<br>**Included outcome measures:**<br>Effect of pelvic floor muscle training on childbirth parameters; effect of perineal massage on childbirth parameters; effect of EPI-NO on childbirth parameters. | **Countries:** Brazil (n=4); Norway (n=4); Canada (n=1); UK (3); Australia (n=2); Germany (n=1); Ireland (n=1); Turkey (n=2); Spain (n=1); France (n=1); Thailand (n=1); no data reported for country and unable to access full text (n=1).<br>**Intervention type:** EPI-NO (n=3); perineal massage (n=6); PFME (n=13)<br>**Delivery method:** N/A for antenatal studies (n=12); normal vaginal delivery (n=5); vaginal and caesarean delivery (n=3) assisted delivery with instruments (n=2). |  | practiced at home during pregnancy. Perineal massage reduced the risk of postpartum perineal pain and could also shorten the second stage of labour. The combined use of interventions that have proven beneficial has not been studied and should be evaluated in the future.<br>Pelvic floor muscle training improved pelvic floor symptoms, and perineal massage improved childbirth-related parameters and pelvic floor symptoms, whereas EPI-NO showed no benefit. |
| Sananès et al. 2023<br>Switzerland | **Review period:** Inception – 9 May 2023<br>**Review purpose:** Umbrella overview of Cochrane systematic reviews of RCTs encompassing antenatal, intrapartum and postpartum interventions for preventing postpartum urinary and faecal incontinence.<br>**Included study designs:** Cochrane systematic reviews.<br>**Included outcome measures:** Not stated. | **Number of included studies:** 9<br>**Key characteristics:**<br>**Study design:** Cochrane systematic reviews (n = 9)<br>**Countries:**<br>**Intervention type:** Caesarean delivery for the prevention of anal incontinence (n = 1), Choice of instruments for assisted vaginal delivery (n = 1), Antenatal perineal massage for reducing perineal trauma (n = 1), Methods of repair for obstetric anal sphincter injury (n = 1), Planned caesarean section for term breech delivery (n = 1), Use of endoanal ultrasound for reducing the risk of complications related to anal sphincter injury after vaginal birth (n = 1), Planned caesarean section for women with a twin pregnancy (n = 1), Selective versus routine use of episiotomy for vaginal birth (n = 1), Pelvic floor muscle training for preventing and treating urinary and faecal<br>incontinence in antenatal and postnatal women (n = 1).<br>**Delivery method:** Caesarean and vaginal birth | **Quality Rating – High (11/11)** | Only the use of a vacuum instead of forceps if an assisted vaginal delivery is needed, the use of an endo-anal ultrasound prior to repairing perineal tears and postpartum pelvic floor muscle training suggest a reduction in postpartum incontinence. Due to the small number of relevant reviews, a consequence of the relatively small number of primary studies, the effect of almost all the tested interventions was found to be imprecise. |
| Santos et al., 2024<br>Brazil | **Review period:** 1966 to September 2022<br>**Review purpose:** To evaluate the effectiveness of aerobic and/or resistance group exercise programs associated with pelvic floor muscle training (PFMT) during prenatal care for the prevention and treatment of urinary incontinence (UI) using the best level of evidence.<br>**Included study designs:** Randomised clinical trials or quasi-randomised.<br>**Included outcome measures:** Self-reported absence of UI, Incontinence-specific quality of life, Self-reported UI, | **Number of included studies:** 5<br>**Key characteristics:**<br>**Study design:** Randomised controlled trials (n=3), quasi randomised (n=2)<br>**Countries:** Norway (n=4), Spain (n=1)<br>**Intervention type:** Aerobic and/or resistance group exercise programs associated with PFMT as an intervention compared to a control group or usual care.<br>**Delivery method:** N/A population was prenatal. | **Quality Rating – High (10/11)** | There was a reduction in the reports of UI postintervention at 16 weeks (RR: 0.83; 95% CI: 0.74–0.93, one study, 762 women, random effects: p = 0.002) and after 3 months (RR: 0.76; 95% CI: 0.60–0.95, one study, 722 women, random effects: p = 0.02), based on moderate certainty of evidence and improvement in UI-specific quality of life (MD: –2.42; 95% CI: –3.32 to –1.52, one study, 151 women, random effects: p < 0.00001), based on low quality of evidence. Other results showed no difference between the postintervention groups, with low and very low evidence.<br>There is moderate evidence that the aerobic and/or resistance exercise program associated with PFMT compared to usual care can reduce postintervention UI, as well as 3 months postintervention, and that it can improve UI-specific quality of life, but with low-evidence certainty. |
| Shinozaki et al., 2022<br>Japan | **Review period:** Inception until January 2021<br>**Review purpose:** this review aimed to systematically assess whether pushing technique used by women in the second stage of labour affects postpartum urinary incontinence and birth outcomes.<br>**Included study designs:** Individual or cluster RCTs.<br>**Included outcome measures:** Primary outcome: UI. Secondary outcomes were perineal related, | **Number of included studies:** 17<br>**Key characteristics:**<br>**Study design:** All n=17 studies were RCTs.<br>**Countries:** USA (n=11), Denmark (n=1), UK (n=1), Iran (n=1), Turkey (n=1), Ireland (n=1) and Canada (n=1).<br>**Intervention type:** Spontaneous pushing<br>**Delivery method:** Vaginal delivery utilising spontaneous pushing (n=6), delayed pushing (n=8), uncoached pushing (n=2) or exhalation while pushing | **Quality Rating – High (10/11)** | The review and meta-analysis results revealed that spontaneous pushing in the second stage of labour statistically reduced the change of scores for UI from baseline compared with directed pushing.<br>After applying the GRADE approach for classifying certainty of evidence, the results showed a very low certainty of evidence for change in UI. Regarding the perineal outcome, the certainties of no suturing and episiotomy were moderate. The duration of the second stage of labour showed a low certainty of evidence. |
|  | such as no suturing of the perineum, third- or fourth-degree laceration and episiotomy, and duration of the second stage of labour. | (n=1). *All of which are classified as spontaneous pushing in this paper.* |  |  |
| Van Kampen et al., 2015<br>Belgium | **Review period:** inception to September 2013.<br>**Review purpose:** to provide a systematic review of clinical studies that investigated the effectiveness of prenatal physiotherapy in treating pregnancy-related symptoms.<br>**Included study designs:** No limit on study design.<br>**Included outcome measures:** Outcome measures were the occurrence, the reduction, the recurrence, or the persistence of pregnancy-related symptoms. | **Number of included studies:** 54<br>**Key characteristics:**<br>**Study design:** RCT (n=36), prospective randomised controlled study (n=12), pilot RCT (n=2), randomised clinical trial (n=2), randomised experimental design (n=2).<br>**Countries:** Norway (n=8), Thailand (n=1), Iran (n=4), Sweden (n=5), South Africa (n=1), USA (n=14), New Zealand (n=1), Canada (n=2), Brazil (n=4), Spain (n=1), Taiwan (n=1), Denmark (n=1), the Netherlands (n=2), Finland (n=1), China (n=1), Germany (n=1), UK (n=2), France (n=1), Switzerland (n=1), Turkey (n=1) and Colombia (n=1).<br>**Intervention type:** For preventing prenatal incontinence: (n=6 PFMT). For treating prenatal incontinence (n=1) PFMT. One study assessed prenatal perineal massage on postnatal incontinence.<br>**Delivery method:** Of n=6 preventing incontinence, n=6 were prenatal. N=1 of postnatal interventions for incontinence involved vaginal delivery. | **Quality Rating – Low (5/11)** | Three studies found a positive effect of antenatal perineal massage in reducing second- or third degree tears or episiotomies Shipman et al. found a stronger effect in the group aged 30 and older Further, Labrecque et al. concluded that massage therapy resulted in less pain 3 months postpartum in women with a previous vaginal birth. Among the pregnant women without previous vaginal birth there were no significant differences between the two groups with regard to perineal pain. Also, the frequencies of dyspareunia and incontinence, for urine, gas or stool, were similar in both groups.<br>Findings on prenatal incontinence (not relevant to review question): Seven randomized controlled trials were included, concerning the effectiveness of PFMT for pregnancy-related UI or FI. The studies can be divided into two groups: a prevention group without incontinence and a treatment group with incontinence at the start of the study. European guidelines already indicated level A evidence, suggesting offering supervised PFMT, lasting at least 3 months, as a first-line therapy to women with stress or mixed incontinence. Two studies found no beneficial effect of PFMT. Possible explanations could be that the training period was not long enough (from 37 weeks' gestation until delivery) or that the standard antenatal care of the control group was sufficient. |
| Wagg & Bunn, 2007<br>UK | **Review period:** from inception to December 2006.<br>**Review purpose:** a report of a systematic review on unassisted pelvic floor exercises for postnatal | **Number of included studies:** 4<br>**Study design:** Randomised controlled trials (n=4); only studies published in peer-reviewed journals were included. | **Quality Rating – High (11/11)** | Unassisted pelvic floor exercises may be helpful in reducing postnatal incontinence, but that effects may not be maintained over time. More high-quality evaluations are needed to establish the efficacy of unassisted pelvic floor treatments and to identify the most appropriate setting for |
|  | stress incontinence and how treatment might be applicable to primary care settings.<br>**Included study designs:**<br>Randomised controlled trials.<br>**Included outcome measures:**<br>pelvic muscle strength, symptoms of incontinence such as leakage on coughing or sneezing, patient satisfaction and quality of life. Unassisted exercises were defined as those where no equipment is used. | **Countries:** Australia (n=1); USA (n=1); UK (n=1); UK and New Zealand (n=1).<br>**Intervention type:** postnatal pelvic floor muscle exercises (n=4).<br>**Delivery method:** Vaginal delivery (n=1), forceps or ventouse delivery only (n=1); vaginal and assisted delivery (n=2). |  | providing this intervention. Research needs to use standardized interventions and outcome measures, include patient relevant outcomes such as quality of life, and have follow-up periods that enable the evaluation of long-term effectiveness.<br>There needs to be more standardization of trials to allow meta-analysis.<br>Unassisted pelvic floor exercises appear to be effective in the short term. Research on long-term effectiveness and use in primary care is needed. |
| Woodley et al., 2020<br>New Zealand | **Review period:** from inception to January 2020.<br>**Review purpose:** To assess the effects of PFMT for preventing or treating urinary and faecal incontinence in pregnant or postnatal women, and summarise the principal findings of relevant economic evaluations.<br>**Included study designs:**<br>Randomised or quasi-randomised trials in which one arm included PFMT.<br>**Included outcome measures:**<br>Prevention of UI; treatment of UI; reduced faecal incontinence; self-reported absence of urinary or faecal incontinence symptoms; Self-reported urinary or faecal incontinence; urinary incontinence-specific quality of life; faecal incontinence-specific quality of life; self-reported severity of incontinence; number of urinary or faecal incontinence episodes; loss | **Number of included studies:** 46<br>**Study design:** Randomised controlled trials (n=41); quasi-randomised trials (n=5).<br>**Countries:** Sweden (n=1); Brazil (n=4); Spain (n=2); Norway (n=5); Australia (n=1); Turkey (n=4); Canada (n=5); UK (n=5); France (n=1); USA (n=3); Italy (n=2); New Zealand (n=2); Mexico (n=1); Republic of Korea (n=1); China (n=5); Switzerland (n=1); Ireland (n=1); Thailand (n=1); Poland (n=1); The Netherlands (n=1).<br>**Intervention type:** PFMT.<br>**Delivery method:** N/A for antenatal studies (n=14); normal vaginal delivery (n=7), vaginal and caesarean delivery (n=6); vaginal, assisted and caesarean delivery (n=9); assisted delivery with instruments (n=3); data not reported in original studies (n=7). | **Quality Rating – High (11/11)** | This review provides evidence that early structured pelvic floor muscle training (PFMT) in early pregnancy for continent women probably prevents the onset of urinary incontinence in late pregnancy and reduces the risk of urinary incontinence slightly postnatally. Population approaches, that is, recruiting antenatal women regardless of their continence status, might also reduce the risk of urinary incontinence in late pregnancy and up to > 3-6 months postpartum, but the effect may be less pronounced. However, the reasons for this are unclear. The findings about the effects of PFMT as a treatment for antenatal urinary incontinence are uncertain. Similarly, it is uncertain whether a population-based approach for delivery of postnatal PFMT (i.e., recruitment of women regardless of continence status immediately following delivery) is effective. It is possible that a 'high-risk' approach (e.g. women who have an assisted delivery or deliver a large baby) leads to more clinical benefit than a population approach. There are insufficient data on faecal incontinence to state whether or not PFMT is effective in preventing or treating this problem in pregnant or postpartum women. Furthermore, there are insufficient data to determine whether |
|  | of urine under stress test; self-reported measures of pelvic floor dysfunction; other self-reported well-being measures; adverse effects, particularly discomfort or pain associated with PFMT; labour and delivery outcomes for women who did antenatal PFMT. |  |  | or not PFMT is effective to prevent urinary incontinence more than one year after birth. |
| Zhang et al., 2023<br>Lead author<br>institution: Spain | **Review period:** 2010 to 2023<br>**Review purpose:** the aim of this systematic review was to examine the effect of PFMT, implemented as a specific section within a physical activity program or as a single intervention during pregnancy, on UI, episiotomy and third- or fourth-degree perineal tear.<br>**Included study designs:** RCTs only. A search for systematic reviews on the topic was conducted to compare results.<br>**Included outcome measures:** The main outcome of the study was UI. Relevant secondary variable outcomes were episiotomy and third- or fourth-degree perineal tear. | **Number of included studies: 30**<br>**Key characteristics:**<br>**Study design:** All (n=30) included studies were RCTs.<br>**Countries:** Malaysia (n=1), Spain (n=6), Norway (n=7), France (n=1), Turkey (n=2), Brazil (n=2), Thailand (n=1), Iran (n=2), China (n=2), Egypt (n=2), Portugal (n=1), Sweden (n=1), Australia (n=1) and England (n=1).<br>**Intervention type:** Exercise programme including PFMT (n=14), yoga including PFMT (n=1), aerobic/strength programme only (n=2), PFMT only (n=9), perineal massage with PFMT (n=4).<br>**Delivery method:** Interventions administered during pregnancy. | **Quality Rating – Moderate (8/11)** | Incorporating regular PFMT or PA including PFMT during pregnancy did not significantly change the outcome in the rate of having an episiotomy.<br>There was an association between the third- or fourth-degree perineal tear and PFMT or PA including PFMT during pregnancy.<br>There was a statistically significant association ($Z = 3.46$; $p < 0.0005$) between PFMT or PA including PFMT during pregnancy and the likelihood of UI. Risk of publication bias testing in the analysed articles showed no potential publication bias ($p = 0.99$) in this analysis.<br>The meta-analyses examining the effects of PFMT on the three study outcomes found a positively significant effect on UI and third- and fourth-degree perineal tears but not on occurrence of episiotomies. The results of this study provide compelling evidence that argues for the promotion and improvement of PFMT among pregnant women without contraindications. |

**Abbreviations:** FI (faecal incontinence); PFM (pelvic floor muscles); PFME (pelvic floor muscle exercises); PFEs (peripartum pelvic floor exercises); PFMT (Pelvic Floor Muscle Training); POP (Pelvic Organ Prolapse); UI (urinary incontinence); SUI (stress urinary incontinence); RCTs (randomised controlled trials); ICS (International Continence Society); QoL (Quality of life).

**Table 3:** Evidence from included economic evaluations (n=3).

| Citation (Country of study) | Study characteristics and health economics methods | Outcomes and costs measured | Quality | Main health economics findings |
| --- | --- | --- | --- | --- |
| Bick et al., 2017<br>England and Wales | **Aim**<br>In nulliparous women with epidural analgesia, does a policy of adopting an 'upright position' throughout second-stage labour increase the incidence of SVB compared with a policy of adopting a 'lying-down' position?<br>**Intervention type**<br>Birthing position: 1) Upright position to maintain the pelvis in as vertical a plane as possible; and<br>(2) lying-down position to maintain the pelvis in as horizontal a plane as possible<br>**Delivery method**<br>Intended spontaneous vaginal delivery following intervention. Data on incidence of instrumental delivery (forceps and ventouse, caesarean section, augmentation of labour) were also collected.<br>**Sample size**<br>3236 women were randomised from 41 centres in England and Wales. Analysed at 1 year follow-up: Upright (n= 950), Lying down (n= 942).<br>**Length of follow-up**<br>1 year<br>**Analytic approach:** trial based economic evaluation.<br>**Type of economic evaluation/cost analysis** | **Outcomes**<br>Incidence of spontaneous vaginal birth (SVB), maternal satisfaction with labour as reported immediately after the birth and urinary and faecal incontinence at 1 year's follow-up. Maternal health-related quality of life was assessed at 1-year follow up using the EQ-5D-3L and SF-12.<br>An Apgar score of < 4 at 5 minutes and major morbidity at 1 year's follow-up were the consequences selected for infants.<br>The secondary cost-effectiveness analysis used the number of additional cases of SVB as the outcome measure.<br>**Types of costs measured**<br>Healthcare service utilization included resources consumed during the late stages of labour to hospital discharge, and during the first 12 months after birth as reported at the 1-year follow-up. No intervention costs as birthing position was not associated with additional resources. | **Quality rating – High (11/11)**<br>JBI critical appraisal checklist for economic evaluations<br>(10 compliant questions, one NA). | **Clinical consequences**<br>The only statistically significant difference observed between trial arms was the higher number of SVBs in the lying-down arm. With 35.2% of women achieving SVB in the upright group, compared with 41.1% in the lying-down group (adjusted relative risk 0.86, 95% CI 0.78 to 0.94). The number of infants in whom cord blood was sampled was slightly higher in the upright group than in the lying-down group, yielding a statistically significant RR (99% CI) of 1.012 (1.010 to 1.013). No other significant differences were detected.<br>There was no evidence of any differences between the groups in relation to the incidence or severity of urinary incontinence, faecal incontinence, constipation, haemorrhoids or dyspareunia, or general well-being. Similarly, there was no evidence of a difference in the incidence of diagnosed cerebral palsy or severe neurodevelopmental delay in any of the infants at 1 year. |
|  | Cost-consequence analysis and secondary cost-effectiveness analysis<br>**Perspective of analysis**<br>NHS perspective<br>**Currency and cost year**<br>British pounds sterling for cost year 2013/14.<br>**Discounting**<br>No discounting as time horizon did not exceed 1 year.<br>**Sensitivity analysis**<br>Sensitivity analysis was conducted which excluded women who had another birth or were pregnant at the time of the 1-year follow-up. | Cost-effectiveness analysis was conducted as a secondary outcome in the economic evaluation. |  | Quality-of-life scores were very similar (almost identical) between trial arms for both instruments and therefore no significant mean differences were detected.<br>**NHS resource use costs**<br>Women randomised to the lying-down position consumed significantly fewer NHS resources than those randomised to an upright position during the original hospital stay [mean cost difference of £59 (95% CI £6 to £111) favouring the lying-down position]. This result was driven by more SVBs in the lying-down arm. At the 12-month follow-up, there were no significant differences in the overall costs incurred by mothers or their babies between the upright and lying-down groups. The significantly higher costs incurred by the women in the upright group were offset by the slightly, but non-significantly, higher costs incurred during follow-up by the women in the lying-down group.<br>**Secondary cost-effectiveness**<br>The ICER (95% CI) was estimated to be £722 (–£2968 to £6358) per additional case of SVB. |
| Brennan et al., 2021<br>Paper is a cost effectiveness analysis of a systematic review; therefore, lead author institution: Australia | **Aim**<br>What is the most cost-effective way of providing pelvic floor muscle training (PFMT) to prevent or treat postpartum incontinence?<br>**Intervention type**<br>Group-based pelvic floor muscle training during pregnancy vs. postnatal training for women with urinary incontinence<br>**Delivery method**<br>Not reported – participants were prenatal or postnatal.<br>**Sample size**<br>Eleven trials were included in the cost-effectiveness analysis.<br>**Length of follow-up**<br>Time points were divided into 0 to 3 months postnatal, >3 to 6 months postnatal and >6 to 12 months postnatal.<br>**Analytic approach:** Economic evaluation using data from a Cochrane systematic review.<br>**Type of economic evaluation/cost analysis**<br>Cost-effectiveness analysis<br>**Perspective of analysis**<br>Health service, consumer and societal perspectives<br>**Currency and cost year**<br>Australian dollars for cost year 2019.<br>**Discounting**<br>NA<br>**Sensitivity analysis** | **Outcomes**<br>Postpartum urinary or faecal incontinence. Sixteen of these trials reported risk ratios for UI, and one further study reported a risk ratio for stress urinary incontinence. Two reported UI-specific quality of life data. Three of the included trials reported risk ratios of self-reported and one reported FI-specific quality of life data. In one study, results were presented for both the Incontinence Impact Questionnaire Short Form (IIQ-7) and the Urogenital Distress Inventory Short Form (UDI- 6), and in this case the results for the IIQ-7 were used.<br>**Types of costs measured**<br>Costs for each model of care were based on publicly available data on costs of consumables, room hire, staffing costs (including hourly rates and on-costs), median hourly wage of women of childbearing age and demographic data on employment levels of women during pregnancy and after childbirth.<br>Consumer costs: out of pocket cost of \$10 per patient was used. Childcare costs were also calculated.<br>Societal costs: The total costs to society were calculated as a sum of the health service costs, consumer costs and the productivity costs that would be caused by women leaving work in order to attend appointments. | **Quality rating - High (11/11)**<br>JBI critical appraisal checklist for economic evaluations<br>(10 compliant questions, one NA). | Three models of care were clinically effective: individually supervised PFMT during pregnancy to prevent urinary incontinence (Model 1), group based PFMT during pregnancy to prevent or treat urinary incontinence (Model 2) and individually supervised postnatal PFMT to treat urinary incontinence and prevent or treat faecal incontinence (Model 3). The health service costs per urinary incontinence case prevented or cured were \$768 for Model 1, and \$1,970 for Model 3. However, Model 2 generated a cost saving of \$14 if there were eight participants per session, with greater savings if more participants attend. The health service cost per faecal incontinence case prevented or cured was \$2,784 (Model 3).<br>Providing group-based PFMT for all women during pregnancy is likely more efficient than individual PFMT for incontinent women postnatally; however, providing PFMT for postnatal women with urinary incontinence should not be discounted because of the added known benefit for preventing and treating faecal incontinence. |
|  | One-way sensitivity analyses to the base case were conducted to account for plausible variations in the number of participants per group for group-based models of care, the cost of patient out of pocket charges, salary rate of the health professional delivering the intervention and the proportion of patients who would have postnatal incontinence without intervention. |  |  |  |
| National Institute of Health and Care Excellence (NICE), 2021b<br>UK<br>(This entry is part of the supporting evidence (Appendix A-R) conducted to inform the NICE guideline [NG210], which covers the prevention, assessment and non-surgical management of pelvic floor dysfunction in women aged 12 and over) | **Aim**<br>As part of the NICE No. 210 Guideline an original economic analysis was undertaken to address the cost-effectiveness of PFMT as a preventative strategy for pelvic floor dysfunction compared to no preventative PFMT in a population of pregnant women. (However, urinary incontinence (UI) was used in the economic evaluation as a proxy for all symptoms of pelvic floor dysfunction.)<br>**Intervention type**<br>Supervised pelvic floor muscle training for the prevention of pelvic floor dysfunction<br>**Delivery method**<br>Delivery method reported in Reilly et al., 2002 and the sample included women who had normal vaginal deliveries, assisted deliveries and caesarean sections.<br>**Sample size**<br>Data was used from two studies reporting on the same intervention at different follow-up time-points.<br>Reilly et al, 2002<br>N = 230, n = 120 assigned to pelvic floor exercise, n = 110 assigned to no intervention. | **Outcomes**<br>The economic analysis focused only on the symptom of urinary incontinence, reflecting the outcomes reported in the randomised controlled trials used to estimate the clinical effectiveness of PFMT.<br>Reilly et al, 2002 – prevalence of incontinence, Pelvic floor muscle strength, Bladder neck mobility, Joint hypermobility, QALY.<br>Agur et al, 2008 - The prevalence of stress urinary incontinence at 8 years.<br>**Types of costs measured**<br>Cost of PFMT session and the monthly health resource costs of urinary incontinence. | **Quality rating (11/11)**<br>JBI critical appraisal checklist for economic evaluations | An intervention to provide antenatal preventative PFMT to prevent pelvic floor dysfunction is cost-effective for a population with a 50% risk at a cost-effectiveness threshold of £20,000 per QALY. The intervention is cost increasing, with a small reduction in the costs of UI management not offsetting the costs of the PFMT sessions. However, the ICER of £11,432 falls below the cost-effectiveness threshold indicating that the QALY gain represents good value for NHS resources. The impact of the transitions between health states over time, indicates that the benefits of preventative PFMT are a consequence of delayed UI onset rather than averted UI.<br>The probabilistic results based on 10,000 model simulations for the base case model and quantify some of the |
|  | Agur et al, 2008<br>Participants were asked about the presence of stress urinary incontinence, impact on quality of life, frequency of performance of PFMT and details of subsequent deliveries. N = 164 women, n = 79 responders received supervised PFMT, n = 85 responders were untreated controls.<br>**Length of follow-up**<br>Reilly et al, 2002 follow up was 28 days, Agur et al, 2008 follow up was 8 years.<br>**Data source of intervention effects:** Two studies identified via systematic review of the literature that reported on the same intervention at different follow-up periods.<br>**Type of economic evaluation/cost analysis**<br>Cost- utility analysis<br>**Perspective of analysis**<br>NHS and Personal Social Services (PSS) perspective<br>**Currency and cost year**<br>Pound sterling for cost year 2020<br>**Discounting**<br>A monthly cost for management of symptoms was applied to women who developed urinary incontinence and an annual discount rate of 3.5%.<br>No discounting was applied to the preventative PFMT intervention as these costs were all incurred within the first year.<br>**Sensitivity analysis**<br>A Dirichlet distribution was used in the probabilistic sensitivity analysis to sample the weighted mean health state utility in women with urinary incontinence. |  |  | uncertainty with respect to the cost-effectiveness conclusions. The mean incremental Net Monetary Benefit of £481 indicates that the preventative PFMT is cost-effective as the value is positive, although there is uncertainty with respect to this conclusion as the 95% credible interval includes negative values. However, preventative PFMT was cost-effective in 63.0% of the simulations. There is a very strong negative correlation between incremental costs and incremental QALYs which is because of the very close link between QALY gains from averted or delayed UI and “downstream” savings from averted or delayed UI.<br>The cost-effectiveness acceptability curve charts the probability that preventative PFMT is cost-effective relative to no PFMT for the base case analysis at different cost-effectiveness thresholds. This indicates that preventative PFMT is likely to be cost-effective relative to no preventative PFMT providing the cost-effectiveness threshold is greater than £11,000 per QALY. It also shows that the probability of preventative PFMT being cost-effective only increases slowly for increasing cost-effectiveness thresholds<br>above £25,000 per QALY. This follows from the non-negligible number of simulations that fall in the north east quadrant of the cost-effectiveness plane where no preventative PFMT dominates (cheaper and more QALYs) preventative PFMT. These simulations would never be considered cost-effective irrespective of the cost-effectiveness threshold. |

**Abbreviations:** EQ-5D-3L (EuroQol-5 Dimensions, three-level version); FI (faecal incontinence); IC (urinary incontinence); SF-12 (Short Form questionnaire-12 items); SVB (spontaneous vaginal birth); ICER (incremental cost-effectiveness ratio).

### 6.3 Quality appraisal

The summary tables for the quality appraisals are presented in Appendix 2.

### 6.4 Information available on request

The data that supports the findings of this study are available in the data extraction tables of this report. The search strategy for Medline via Ovid is available in Appendix 1.

## Acknowledgements

The authors would like to thank Janine Hale, Anjana Kaur and Beti-Jane Ingram for their time, expertise, and contributions during stakeholder meetings in guiding the focus of the review and interpretation of findings. We would also like to thank Elizabeth Gillen and Juliette Hounsome, Information Specialists at Cardiff University for their expert contribution to the search strategy development and database searches.

## 8. APPENDIX

## 8.1 APPENDIX 1: Search strategy for Medline via Ovid

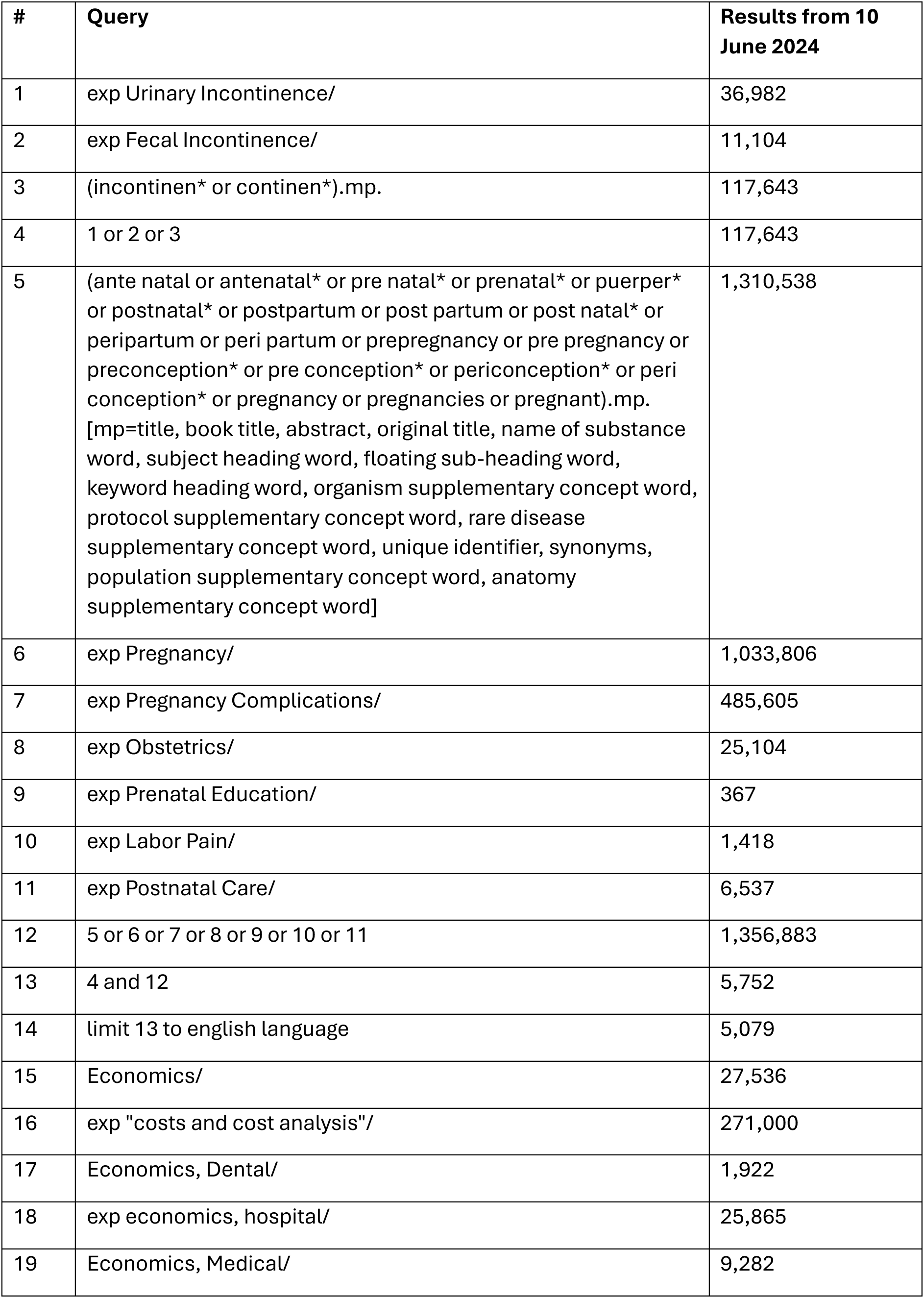
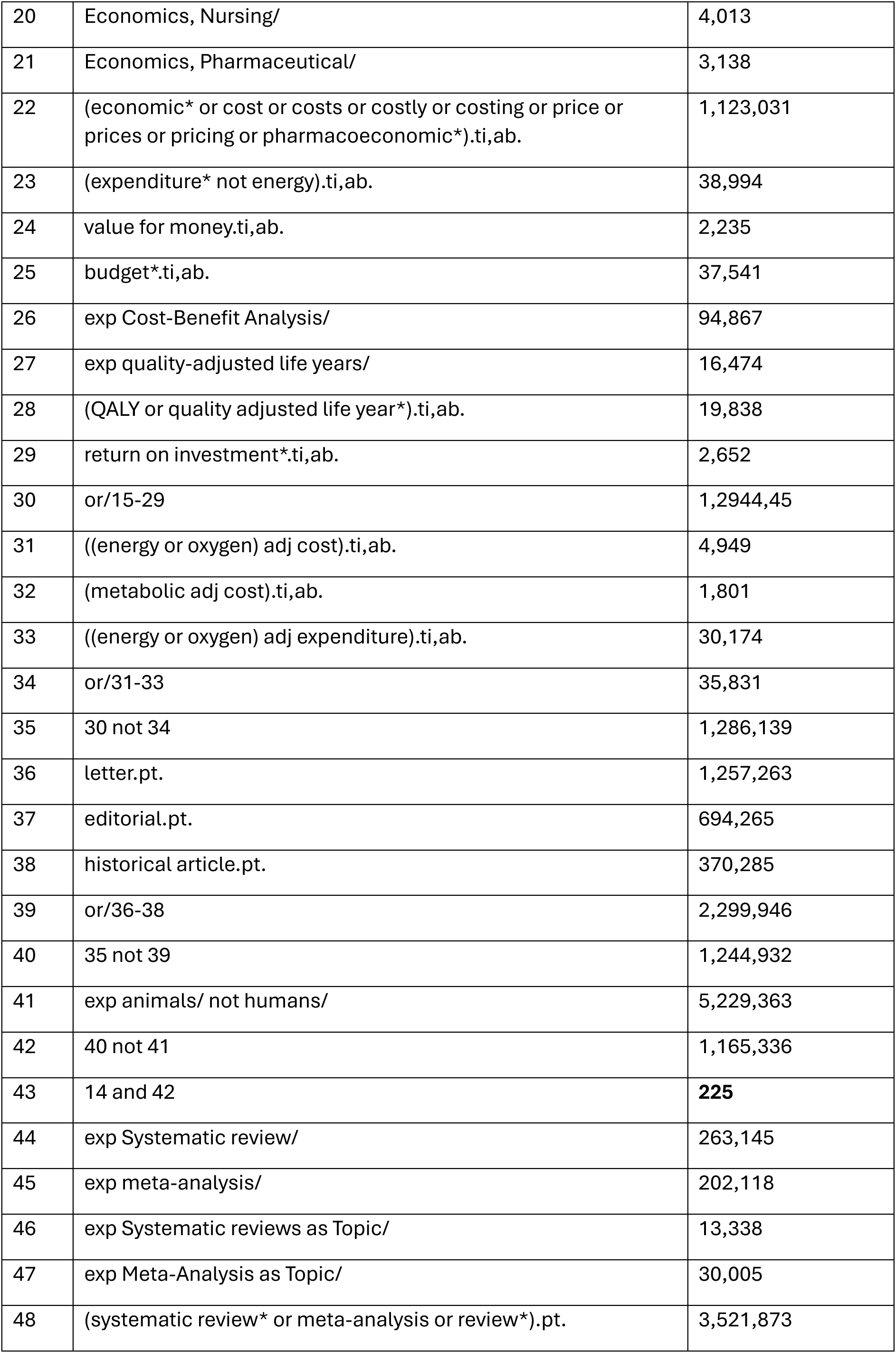
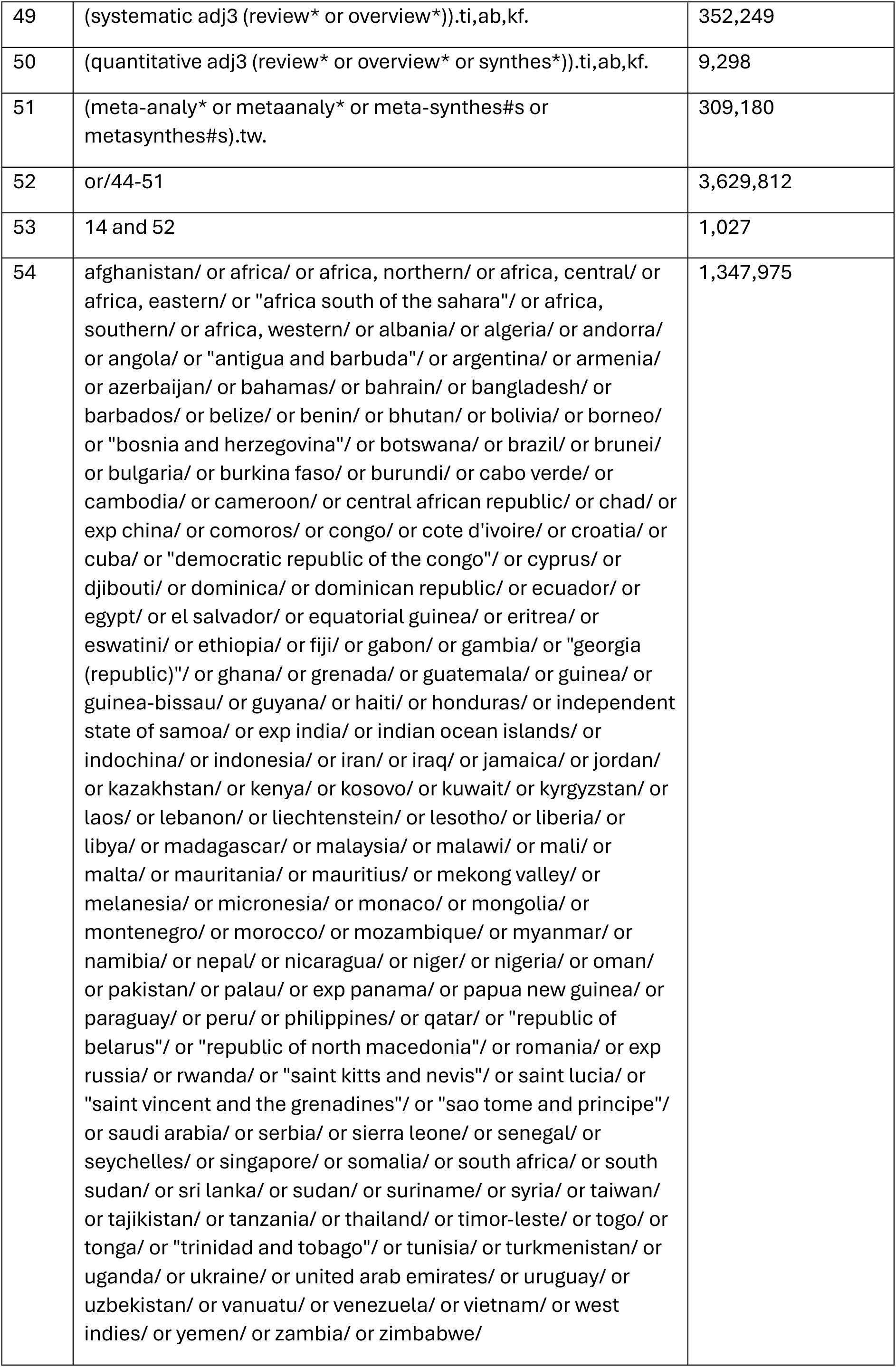
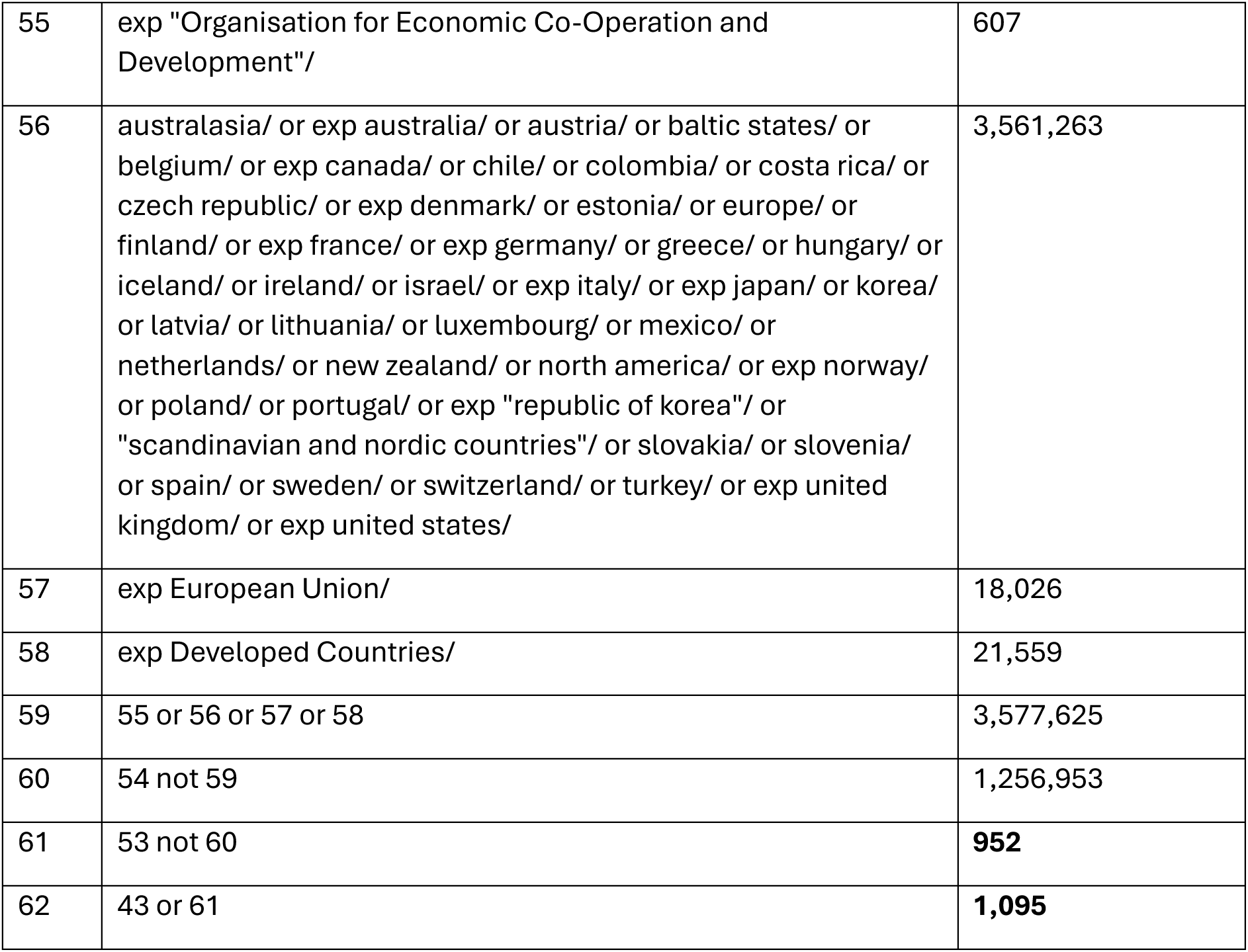

## 8.2 APPENDIX 2: Quality appraisal tables

### JBI critical appraisal of systematic reviews

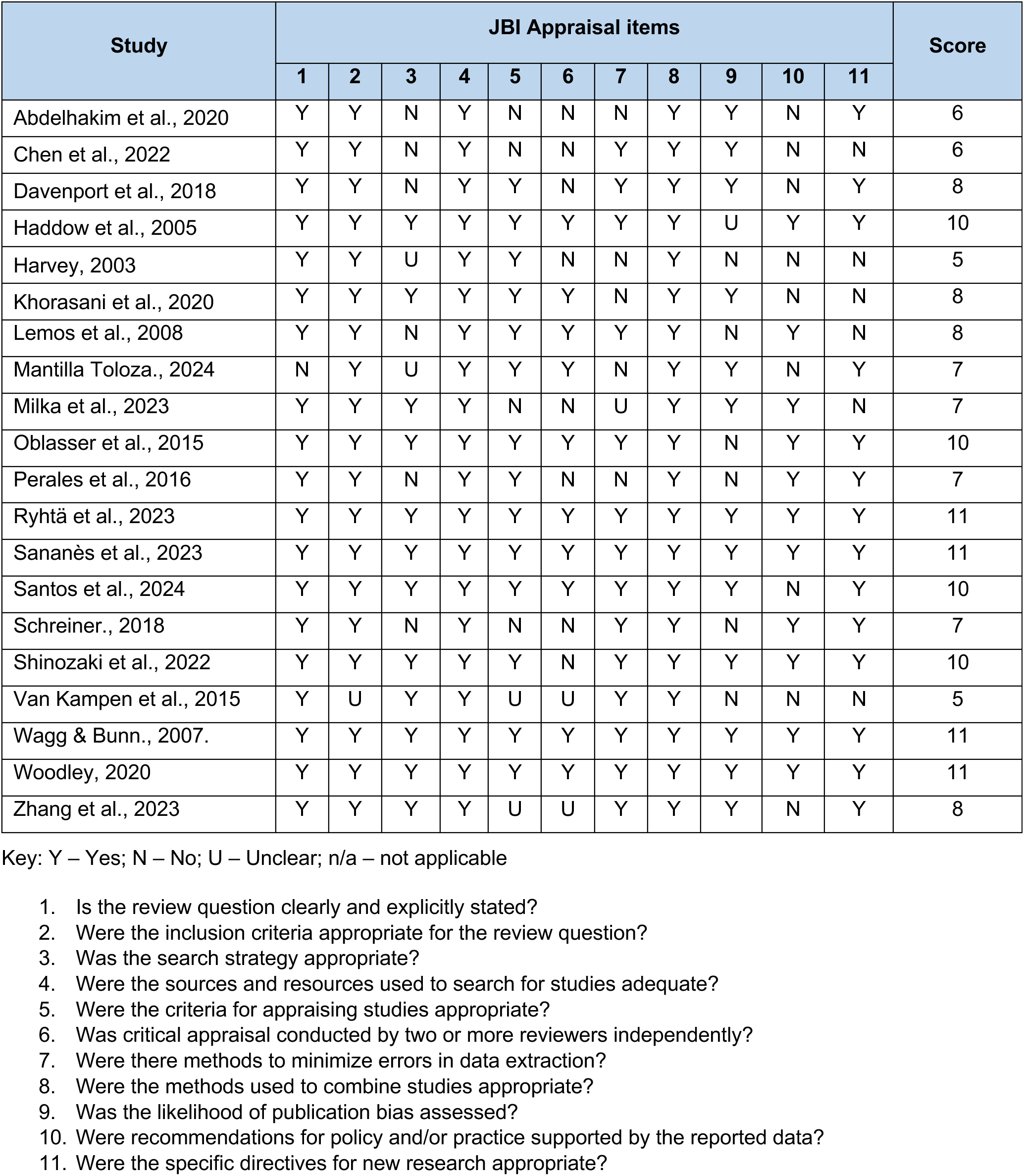

8.2 APPENDIX 2: Quality appraisal tables JBİ critical appraisal of systematic reviews
| Study | JBİ Appraisal items |  |  |  |  |  |  |  |  |  |  | Score |
| --- | --- | --- | --- | --- | --- | --- | --- | --- | --- | --- | --- | --- |
|  | 1 | 2 | 3 | 4 | 5 | 6 | 7 | 8 | 9 | 10 | 11 |  |
| Abdelhakim et al., 2020 | Y | Y | N | Y | N | N | N | Y | Y | N | Y | 6 |
| Chen et al., 2022 | Y | Y | N | Y | N | N | Y | Y | Y | N | N | 6 |
| Davenport et al., 2018 | Y | Y | N | Y | Y | N | Y | Y | Y | N | Y | 8 |
| Haddow et al., 2005 | Y | Y | Y | Y | Y | Y | Y | Y | U | Y | Y | 10 |
| Harvey, 2003 | Y | Y | U | Y | Y | N | N | Y | N | N | N | 5 |
| Khorasani et al., 2020 | Y | Y | Y | Y | Y | Y | N | Y | Y | N | N | 8 |
| Lemos et al., 2008 | Y | Y | N | Y | Y | Y | Y | Y | N | Y | N | 8 |
| Mantilla Toloza., 2024 | N | Y | U | Y | Y | Y | N | Y | Y | N | Y | 7 |
| Milka et al., 2023 | Y | Y | Y | Y | N | N | U | Y | Y | Y | N | 7 |
| Oblasser et al., 2015 | Y | Y | Y | Y | Y | Y | Y | Y | N | Y | Y | 10 |
| Perales et al., 2016 | Y | Y | N | Y | Y | N | N | Y | N | Y | Y | 7 |
| Ryhtä et al., 2023 | Y | Y | Y | Y | Y | Y | Y | Y | Y | Y | Y | 11 |
| Sananès et al., 2023 | Y | Y | Y | Y | Y | Y | Y | Y | Y | Y | Y | 11 |
| Santos et al., 2024 | Y | Y | Y | Y | Y | Y | Y | Y | Y | N | Y | 10 |
| Schreiner., 2018 | Y | Y | N | Y | N | N | Y | Y | N | Y | Y | 7 |
| Shinozaki et al., 2022 | Y | Y | Y | Y | Y | N | Y | Y | Y | Y | Y | 10 |
| Van Kampen et al., 2015 | Y | U | Y | Y | U | U | Y | Y | N | N | N | 5 |
| Wagg & Bunn., 2007. | Y | Y | Y | Y | Y | Y | Y | Y | Y | Y | Y | 11 |
| Woodley, 2020 | Y | Y | Y | Y | Y | Y | Y | Y | Y | Y | Y | 11 |
| Zhang et al., 2023 | Y | Y | Y | Y | U | U | Y | Y | Y | N | Y | 8 |

Key: Y – Yes; N – No; U – Unclear; n/a – not applicable
1. Is the review question clearly and explicitly stated? 2. Were the inclusion criteria appropriate for the review question? 3. Was the search strategy appropriate? 4. Were the sources and resources used to search for studies adequate? 5. Were the criteria for appraising studies appropriate? 6. Was critical appraisal conducted by two or more reviewers independently? 7. Were there methods to minimize errors in data extraction? 8. Were the methods used to combine studies appropriate? 9. Was the likelihood of publication bias assessed? 10. Were recommendations for policy and/or practice supported by the reported data? 11. Were the specific directives for new research appropriate?

### JBI CRITICAL APPRAISAL CHECKLIST FOR ECONOMIC EVALUATIONS

| Citation | Q1 Is there a well-defined question? | Q2 Is there a comprehensive description of alternatives? | Q3 Are all important and relevant costs and outcomes for each alternative identified? | Q4 Has clinical effectiveness been established? | Q5 Are costs and outcomes measured accurately? | Q6 Are costs and outcomes valued credibly? | Q7 Are costs and outcomes adjusted for differential timing? | Q8 Is there an incremental analysis of costs and consequences? | Q9 Were sensitivity analyses conducted to investigate uncertainty in estimates of cost or consequences? | Q10 Do study results include all issues of concern to users? | Q11 Are the results generalizable to the setting of interest in the review? |
| --- | --- | --- | --- | --- | --- | --- | --- | --- | --- | --- | --- |
| Bick et al., 2017 | Y | Y | Y | Y | Y | Y | NA | Y | Y | Y | Y |
| Brennan et al., 2021 | Y | Y | Y | Y | Y | Y | NA | Y | Y | Y | Y |
| NICE, 2021b | Y | Y | Y | Y | Y | Y | Y | Y | Y | Y | Y |

Y = Yes; N = No; NA = not applicable; U = unclear

## 8.3 APPENDIX 3: Mapping of outcomes reported in the systematic review papers

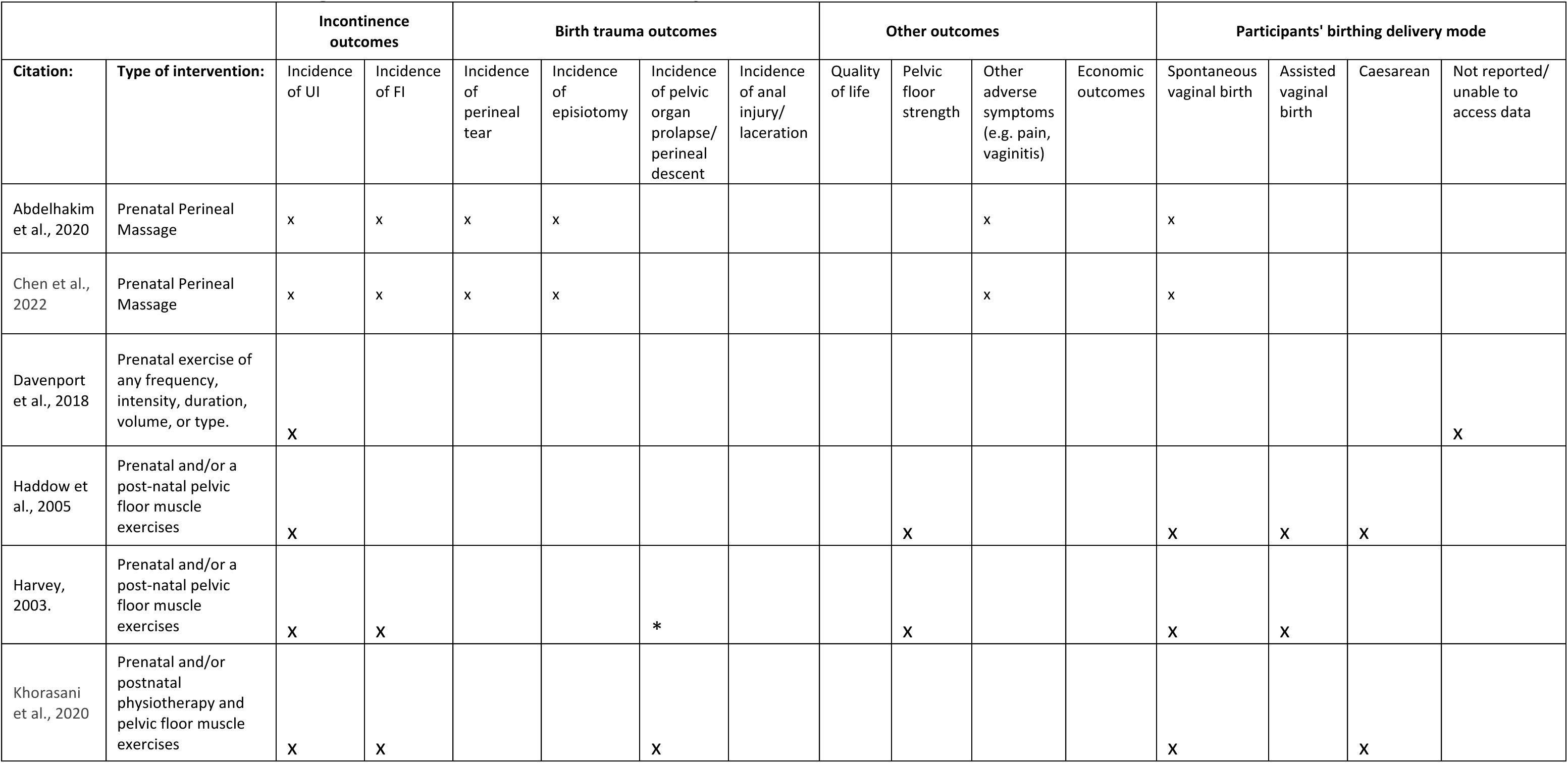
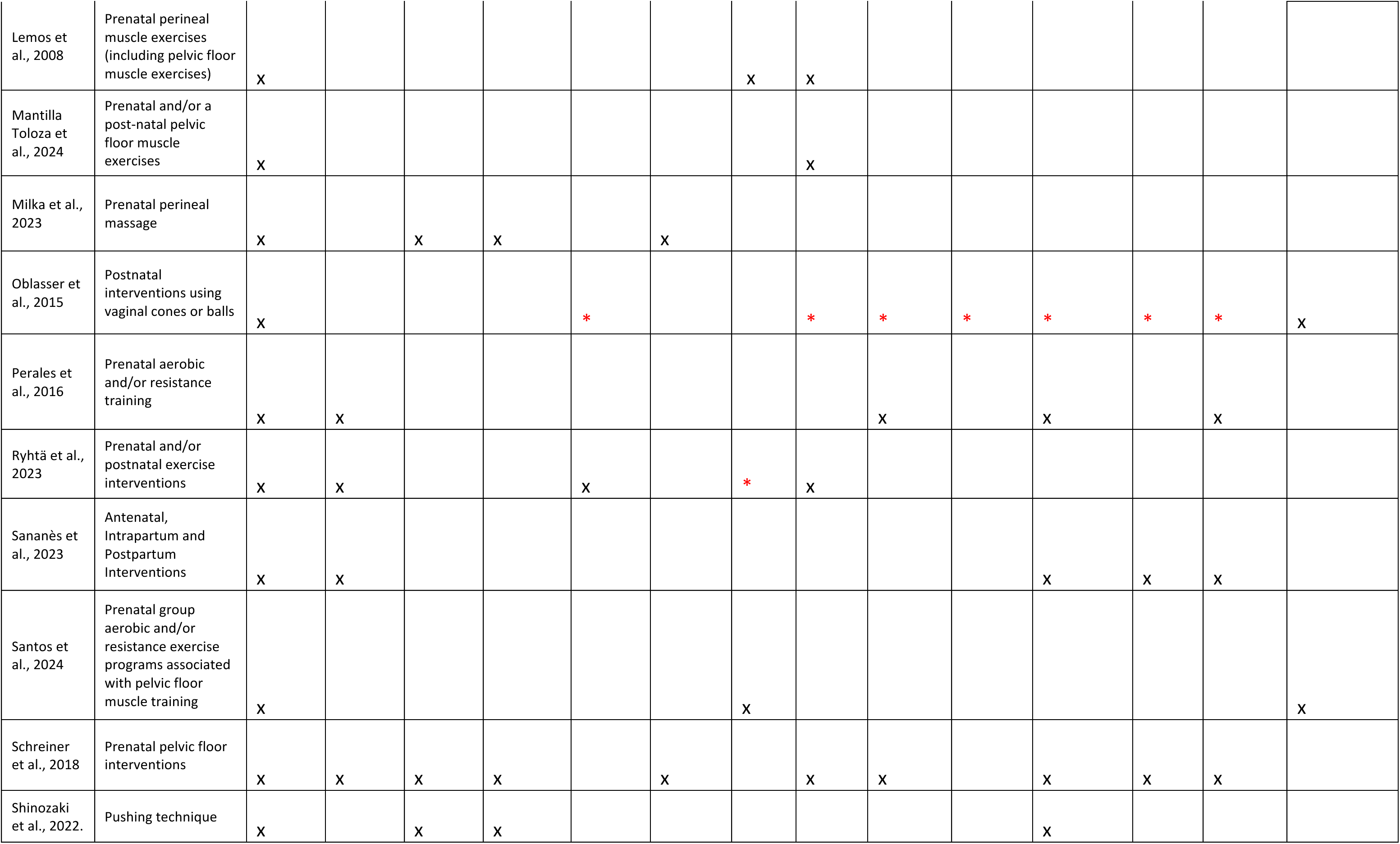
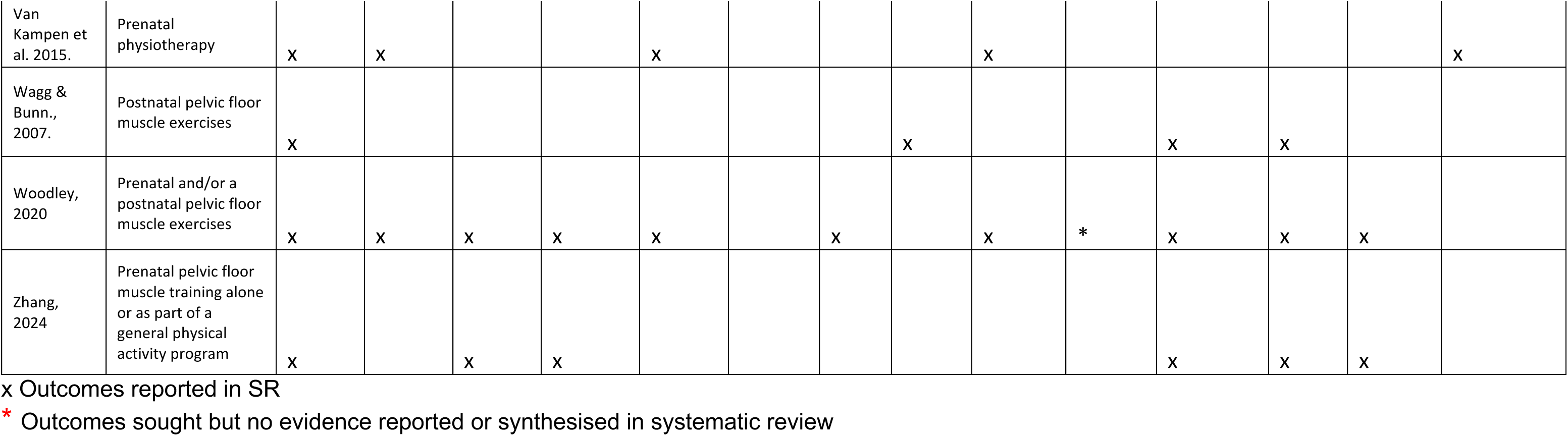

## 8.4 APPENDIX 4: Graphical Representation of Overlap for OVErviews (GROOVE) (Bracchiglione et al., 2022)

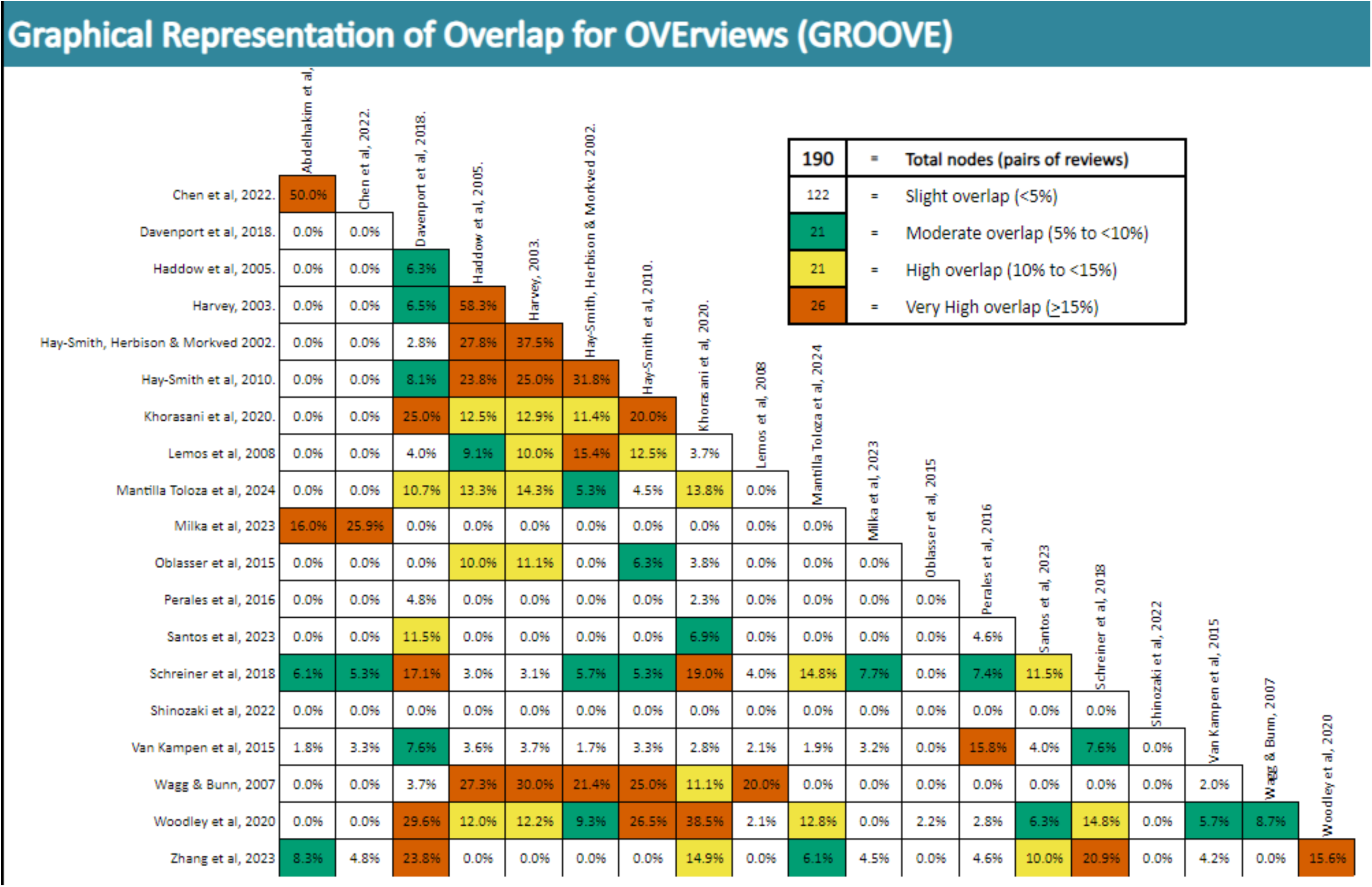
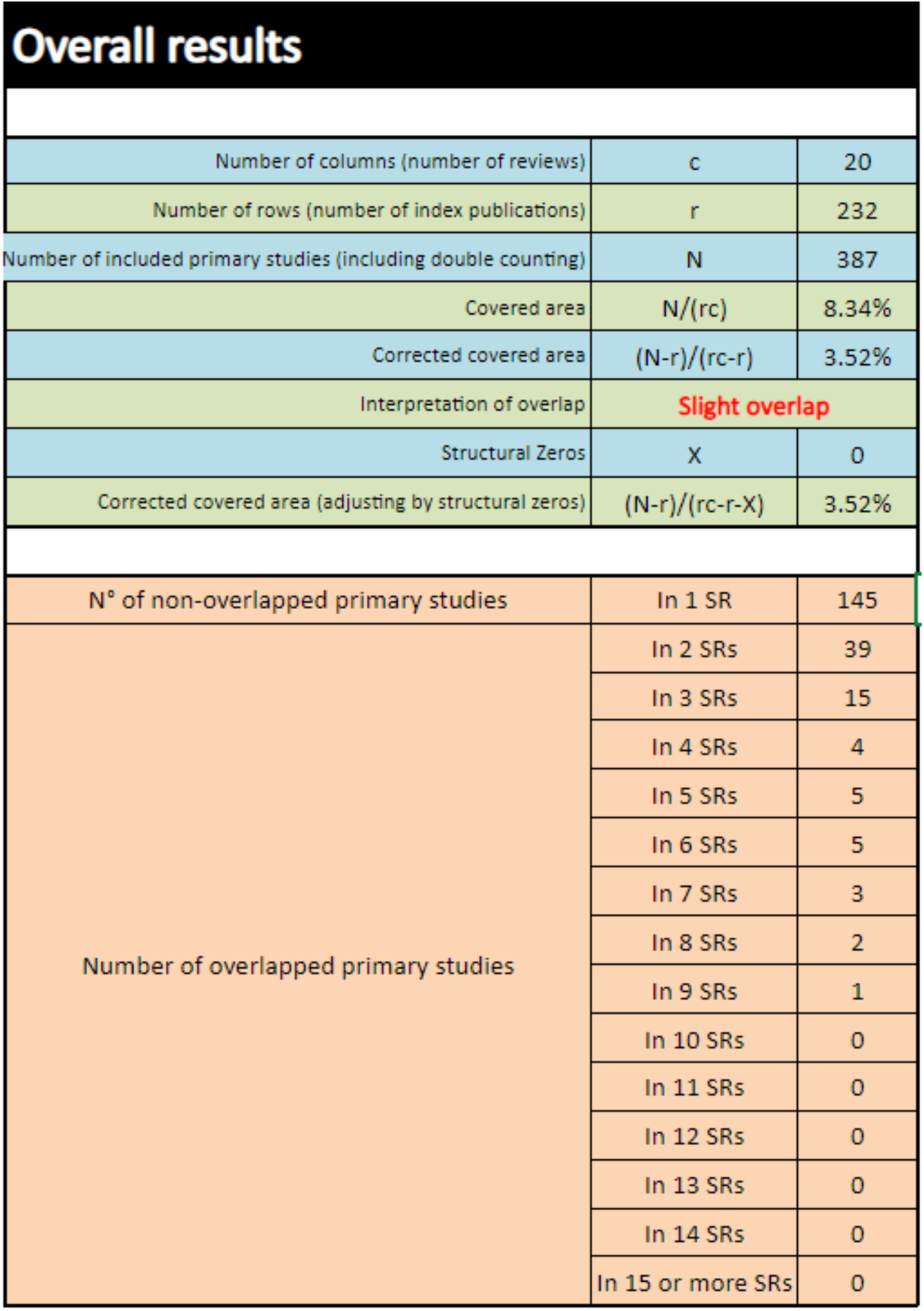

## The Health and Care Research Wales Evidence Centre

Our dedicated team works together with Welsh Government, the NHS, social care, research institutions and the public to deliver vital research to tackle health and social care challenges facing Wales.

Funded by Welsh Government, through Health and Care Research Wales, the Evidence Centre answers key questions to improve health and social care policy and provision across Wales.

Along with our collaborating partners, we conduct reviews of existing evidence and new research, to inform policy and practice needs, with a focus on ensuring real-world impact and public benefit that reaches everyone.

## REFERENCES

Agur, W., Steggles, P., Waterfield, M., & Freeman, R. (2008). Does antenatal pelvic floor muscle training affect the outcome of labour? A randomised controlled trial. International urogynecology journal and pelvic floor dysfunction, 19(1), 85–88. 10.1007/s00192-007-0391-z

Aluko P, Graybill E, Craig D, Henderson C, Drummond M, Wilson ECF, Robalino S, Vale L; on behalf of the Campbell and Cochrane Economics Methods Group. (2023). Chapter 20: Economic evidence. In: Higgins JPT, Thomas J, Chandler J, Cumpston M, Li T, Page MJ, Welch VA (editors). Cochrane Handbook for Systematic Reviews of Interventions version 6.4 (updated August 2023). Available from www.training.cochrane.org/handbook.

Ayiku L, Hudson T, Williams C, Levay P, Jacob C. (2021). The NICE OECD countries’ geographic search filters: Part 2-validation of the MEDLINE and Embase (Ovid) filters. J Med Libr Assoc.1;109(4):583–589. 10.5195/jmla.2021.1224.

Bauer A, Tinelli M and Knapp M (2022). The Economic Case for Increasing Access to Treatment for Women with Common Mental Health Problems During the Perinatal Period, Care Policy and Evaluation Centre, London. https://www.lse.ac.uk/cpec/assets/documents/CPEC-Perinatal-Economics-2022.pdf

Bick, D., Briley, A., Brocklehurst, P., Hardy, P., Juszczak, E., Lynch, L., … & Wilson, M. (2017). A multicentre, randomised controlled trial of position during the late stages of labour in nulliparous women with an epidural: clinical effectiveness and an economic evaluation (BUMPES). Health Technology Assessment (Winchester, England), 21(65), 1. 10.3310/hta21650

Bracchiglione, J., Meza, N., Bangdiwala, S. I., Niño de Guzmán, E., Urrútia, G., Bonfill, X., & Madrid, E. (2022). Graphical representation of overlap for overviews: GROOVE tool. Research Synthesis Methods, 13(3), 381–388. Graphical representation of overlap for overviews: GROOVE tool. Research Synthesis Methods, 13(3), pp.381–388. 10.1002/jrsm.1557

Beckmann MM, Stock OM. (2013). Antenatal perineal massage for reducing perineal trauma. Cochrane Database Syst Rev. (4):CD005123. 10.1002/14651858.CD005123.pub3

Brandie, M., Mackenzie, A. (2009). Perineal trauma following vaginal delivery. Journal of the Association of Chartered Physiotherapists in Women’s Health. 105, 40–55.

Brennen R, Frawley HC, Martin J, Haines TP. (2021). Group-based pelvic floor muscle training for all women during pregnancy is more cost-effective than postnatal training for women with urinary incontinence: cost-effectiveness analysis of a systematic review. J Physiother. 67(2):105–114.

Chen Y, Geng X, Zhou H, Wang W, Liang Y, Zhang C, Wang L. (2022). Systematic review and meta-analysis of evaluation of selective cesarean section in postpartum pelvic floor function recovery under perineal ultrasound. Ann Palliat Med. 11(2):730–742. 10.21037/apm-22-46

Davenport, M. H., Nagpal, T. S., Mottola, M. F., Skow, R. J., Riske, L., Poitras, V. J., … & Ruchat, S. M. (2018). Prenatal exercise (including but not limited to pelvic floor muscle training) and urinary incontinence during and following pregnancy: a systematic review and meta-analysis. British journal of sports medicine, 52(21), 1397–1404. 10.1136/bjsports-2018-099780

Fritel, X., Ringa, V., Quiboeuf, E., & Fauconnier, A. (2012). Female urinary incontinence, from pregnancy to menopause: a review of epidemiological and pathophysiological findings. Acta obstetricia et gynecologica Scandinavica, 91(8), 901–910.v 10.1111/j.1600-0412.2012.01419.x

Fultz, N., Girts, T., Kinchen, K., Nygaard, I., Pohl, G., Sternfeld, B. (2005). Prevalence, management and impact of urinary incontinence in the workplace, Occupational Medicine, 55 (7), 552–557, 10.1093/occmed/kqi152

Grant, A., Currie, S. (2020). Qualitative exploration of the acceptability of a postnatal pelvic floor muscle training intervention to prevent urinary incontinence. BMC Women’s Health, 20, 9. 10.1186/s12905-019-0878-z

Haddow, G., Watts, R., & Robertson, J. (2005). Effectiveness of a pelvic floor muscle exercise program on urinary incontinence following childbirth. JBI library of systematic reviews, 3(5), 1–62. 10.11124/01938924-200503050-00001

Harvey M. A. (2003). Pelvic floor exercises during and after pregnancy: a systematic review of their role in preventing pelvic floor dysfunction. Journal of obstetrics and gynaecology Canada: JOGC = Journal d’obstetrique et gynecologie du Canada: JOGC, 25(6), 487–498.

Javanbakht, M., Moloney, E., Brazzelli, M., Wallace, S., Ternent, L., Omar, M. I., … & Craig, D. (2020). Economic evaluation of surgical treatments for women with stress urinary incontinence: a cost-utility and value of information analysis. BMJ open, 10(6), e035555. 10.1136/bmjopen-2019-035555

Joanna Briggs Institute. (2017a). Checklist for Economic Evaluations. Available at: https://jbi.global/sites/default/files/2019-05/JBI_Critical_Appraisal-Checklist_for_Economic_Evaluations2017_0.pdf

Joanna Briggs Institute. (2017b). Checklist for Systematic Reviews and Research Syntheses. Available at: http://joannabriggs.org/research/critical-appraisal-tools.html

Jones K, Webb S, Manresa M, Hodgetts-Morton V, Morris RK. (2019). The incidence of wound infection and dehiscence following childbirth-related perineal trauma: A systematic review of the evidence. Eur J Obstet Gynecol Reprod Biol. 240:1–8. 10.1016/j.ejogrb.2019.05.038.

Khorasani F, Ghaderi F, Sarbakhsh P, Ahadi P, Khorasani, E, Ansari, F, Vahed, N. (2019). Physiotherapy and Pelvic Floor Muscle Exercises for the Prevention and Treatment of Pregnancy-Related Pelvic Floor Dysfunctions: A Systematic Review and Meta-analysis. International Journal of Women’s Health and Reproduction Sciences. 8. 125–132. 10.15296/ijwhr.2020.20.

Lemos, A., de Souza, A. I., Ferreira, A. L. C. G., Figueiroa, J. N., & Cabral-Filho, J. E. (2008). Do perineal exercises during pregnancy prevent the development of urinary incontinence? A systematic review. International journal of urology, 15(10), 875–880. 10.1111/j.1442-2042.2008.02145.x

Mantilla Toloza, S., Villareal Cogollo, A., & Peña García, K. (2024). Pelvic floor training to prevent stress urinary incontinence: a systematic review. Actas Urológicas Españolas (English Edition), 48(4), 319–327. 10.1016/j.acuroe.2024.01.007

Marchisio S, Ferraccioli K, Barbieri A, Porcelli A, Panella M. Care pathways in obstetrics: the effectiveness in reducing the incidence of episiotomy in childbirth. J Nurs Manag. 2006 Oct;14(7):538–43. 10.1111/j.1365-2934.2006.00704.x. PMID: 17004964.

Menezes, M., Pereira, M., & Hextall, A. (2010). Predictors of female urinary incontinence at midlife and beyond. Maturitas, 65(2), 167–171. 10.1016/j.maturitas.2009.10.004

Milka W, Paradowska W, Kołomańska-Bogucka D, Mazur-Bialy AI. (2023). Antenatal perineal massage - risk of perineal injuries, pain, urinary incontinence and dyspereunia - a systematic review. J Gynecol Obstet Hum Reprod. 52(8):102627. 10.1016/j.jogoh.2023.102627

Moossdorff-Steinhauser H.F.A, Berghmans B.C.M, Spaanderman M.E.A, Bols E.M.J. (2021). Prevalence, incidence and bothersomeness of urinary incontinence in pregnancy: a systematic review and meta-analysis. Int Urogynecol. 32(7):1633–1652. 10.1007/s00192-020-04636-3

Moran PS, Wuytack F, Turner M, Normand C, Brown S, Begley C, Daly D. (2020). Economic burden of maternal morbidity - A systematic review of cost-of-illness studies. PLoS One.16;15(1):e0227377. 10.1371/journal.pone.0227377

NHS (2019). The NHS Long Term Plan. Available at: https://www.longtermplan.nhs.uk/wp-content/uploads/2019/08/nhs-long-term-plan-version-1.2.pdf

National Health Service Digital. (2019). NHS maternity statistics 2018-2019. Available at: https://files.digital.nhs.uk/D0/C26F84/hosp-epis-stat-mat-summaryreport-2018-19.pdf

National Institute for Health and Care Excellence (NICE). (2021a). Pelvic floor dysfunction: prevention and non-surgical management. Available at https://www.ncbi.nlm.nih.gov/books/NBK579556/

National Institute for Health and Care Excellence (NICE). (2021b). Pelvic floor dysfunction: prevention and non-surgical management [F] Pelvic floor muscle training for the prevention of pelvic floor dysfunction evidence review. Available at: https://www.ncbi.nlm.nih.gov/books/NBK579553/

National Institute for Health and Care Excellence (NICE). (2021c). Urinary incontinence in Women [QS77]. Available at: https://www.nice.org.uk/guidance/qs77/resources/urinary-incontinence-in-women-pdf-2098853147077

National Institute for Health and Care Excellence (2022). NICE health technology evaluations: the manual. Process and methods [PMG36]. Available at: https://www.nice.org.uk/process/pmg36

National Institute for Health and Care Excellence (2023). Intrapartum care for health women and babies [I] Interventions to reduce perineal trauma [CG190]. Available at: https://www.nice.org.uk/guidance/ng235/documents/evidence-review-9

Oblasser C, Christie J, McCourt C. (2015). Vaginal cones or balls to improve pelvic floor muscle performance and urinary continence in women post partum: A quantitative systematic review. Midwifery. 31(11):1017–25. 10.1016/j.midw.2015.08.011

Orlovic, M., Carter, A.W., Marti, J., Mossialos, E. (2016). Estimating the incidence and the economic burden of third and fourth-degree obstetric tears in the English NHS: an observational study using propensity score matching. BMJ Open,12;7(6). 10.1136/bmjopen-2016-015463

Okeahialam, N.A., Sultan, A.H., Thakar, R. (2022). The prevention of perineal tramua during vaginal birth. The American Journal of Obstetrics and Gyneacology. 10.1016/j.ajog.2022.06.021

Page MJ, McKenzie JE, Bossuyt PM, Boutron I, Hoffmann TC, Mulrow CD, Shamseer L, Tetzlaff JM, Akl EA, Brennan SE, Chou R, Glanville J, Grimshaw JM, Hróbjartsson A, Lalu MM, Li T, Loder EW, Mayo-Wilson E, McDonald S, McGuinness LA, Stewart LA, Thomas J, Tricco AC, Welch VA, Whiting P, Moher D. The PRISMA 2020 statement: an updated guideline for reporting systematic reviews. (2021). BMJ. 29;372:n71. 10.1136/bmj.n71.

Papanicolaou, S., Pons, M. E., Hampel, C., Monz, B., Quail, D., von der Schulenburg, M. G., … & Sykes, D. (2005). Medical resource utilisation and cost of care for women seeking treatment for urinary incontinence in an outpatient setting: examples from three countries participating in the PURE study. Maturitas, 52, 35–47. 10.1016/j.maturitas.2005.09.004

Perales, M., Santos-Lozano, A., Ruiz, J. R., Lucia, A., & Barakat, R. (2016). Benefits of aerobic or resistance training during pregnancy on maternal health and perinatal outcomes: A systematic review. Early human development, 94, 43–48. 10.1016/j.earlhumdev.2016.01.004

Reilly, E. T. C., Freeman, R. M., Waterfield, M. R., Waterfield, A. E., Steggles, P., & Pedlar, F. (2002). Prevention of postpartum stress incontinence in primigravidae with increased bladder neck mobility: a randomised controlled trial of antenatal pelvic floor exercises. BJOG: An International Journal of Obstetrics and Gynaecology, 109(1), 68–76. 10.1111/j.1471-0528.2002.t01-1-01116.x

Royal College of Obstetricians and Gynaecologists (n.d.) Perineal tears during childbirth. Available: https://www.rcog.org.uk/for-the-public/perineal-tears-and-episiotomies-in-childbirth/perineal-tears-during-childbirth/

Royal College of Obstetricians and Gynaecologists (2015). The Management of Third- and Fourth-Degree Perineal Tears. Green-top Guideline No. 29. Available at: https://www.missionmrcog.com/home/images/Library/Greentop_Guidelines/2015/002_GTG_29_THRID_AND_FORTH_DEGREE_TEAR.pdf

Ryhtä, I., Axelin, A., Parisod, H., Holopainen, A., & Hamari, L. (2023). Effectiveness of exercise interventions on urinary incontinence and pelvic organ prolapse in pregnant and postpartum women: umbrella review and clinical guideline development. JBI Evidence Implementation, 21(4), 394–408. 10.1097/XEB.0000000000000391

Sananès, J., Pire, S., Feki, A., Boulvain, M., & Faltin, D. L. (2023). Antenatal, Intrapartum and Postpartum Interventions for Preventing Postpartum Urinary and Faecal Incontinence: An Umbrella Overview of Cochrane Systematic Reviews. Journal of clinical medicine, 12(18), 6037. 10.3390/jcm12186037

Santos AC, Dias SN, Delgado A, Lemos A. (2024). Effectiveness of group aerobic and/or resistance exercise programs associated with pelvic floor muscle training during prenatal care for the prevention and treatment of urinary incontinence: A systematic review. Neurourol Urodyn. 43(1):205–218. 10.1002/nau.25309

Schreiner, L., Crivelatti, I., de Oliveira, J. M., Nygaard, C. C., & Dos Santos, T. G. (2018). Systematic review of pelvic floor interventions during pregnancy. International journal of gynaecology and obstetrics: the official organ of the International Federation of Gynaecology and Obstetrics, 143(1), 10–18. 10.1002/ijgo.12513

Smith LA, Price N, Simonite V, Burns EE. (2013). Incidence of and risk factors for perineal trauma: a prospective observational study. BMC Pregnancy Childbirth. 13:59. 10.1186/1471-2393-13-59.

Sobhgol SS, Smith CA, Thomson R, Dahlen HG. (2022). The effect of antenatal pelvic floor muscle exercise on sexual function and labour and birth outcomes: A randomised controlled trial. Women Birth. 35(6):e607–e614. 10.1016/j.wombi.2022.02.009.

Steen, M., Diaz, M. (2018). Perineal trauma: A women’s health and wellbeing issue. British Journal of Midwifery, 26 (9), 574-584. 10.12968/bjom.2018.26.9.574

The All-Party Parliamentary Group on Birth Trauma. (2024). Listen to Mums: Ending the Postcode Lottery on Perinatal Care. Available: https://www.theo-clarke.org.uk/sites/www.theo-clarke.org.uk/files/2024-05/Birth%20Trauma%20Inquiry%20Report%20for%20Publication_May13_2024.pdf

Thom, D.H., Rortveit, G. (2010). Prevalence of postpartum urinary incontinence: a systematic review. Acta Obstetrica et Gynecologica Scandinavia, 89 (12), 1511- 1522. 10.3109/00016349.2010.526188

Turner, D. A., Shaw, C., McGrother, C. W., Dallosso, H. M., & Cooper, N. J. (2004). The cost of clinically significant urinary storage symptoms for community dwelling adults in the UK. BJU international, 93(9), 1246–1252. 10.1111/j.1464-410x.2004.04806.x

UK Government (2020). First Do No Harm – The report of the Independent Medicines and Medical Devices Safety Review. Available at: https://www.immdsreview.org.uk/downloads/IMMDSReview_Web.pdf

Van Kampen, M., Devoogdt, N., De Groef, A., Gielen, A., & Geraerts, I. (2015). The efficacy of physiotherapy for the prevention and treatment of prenatal symptoms: a systematic review. International urogynecology journal, 26, 1575–1586. 10.1007/s00192-015-2684-y

Wagg A, Bunn F. (2007). Unassisted pelvic floor exercises for postnatal women: a systematic review. J Adv Nurs. 58(5):407–17. 10.1111/j.1365-2648.2007.04318.x

Wilson, D., Dornan, J., Milsom, I., & Freeman, R. (2014). UR-CHOICE: can we provide mothers-to-be with information about the risk of future pelvic floor dysfunction? International Urogynecology Journal, 25(11), 1449–1452. 10.1007/S00192-014-2376-Z/METRICS

Woodley, S. J., Lawrenson, P., Boyle, R., Cody, J. D., Mørkved, S., Kernohan, A., … & Hay-Smith, J. (2020). Pelvic floor muscle training for preventing and treating urinary and faecal incontinence in antenatal and postnatal women. Cochrane Database of Systematic Reviews, 2021(3). 10.1002/14651858.cd007471.pub4

Zhang X, Mai Y, Hu X. (2023). Effect of pelvic floor muscle training on pelvic floor muscle function and lower urinary tract symptoms in stroke patients: a systematic review. Physiother Theory Pract. 3;39(7):1342-1354. 10.1080/09593985.2022.2040668

